# Disrupted O-GlcNAc homeostasis links microglial dysfunction to cerebellar hypoplasia

**DOI:** 10.64898/2026.09.26.26363519

**Authors:** Sérgio Almeida, Shagana T Balasubramaniam, Anastazja Jusiel, Tim-Simon Burmeister, Leonor Martins, Gitte Ulbjerg Toft, Carolina Lebre, Katrin Õunap, Pilvi Ilves, Lin Lin, Marina Romero-Ramos, João Peça, Joana Guedes, Daan M.F. van Aalten

**Author notes:** These authors contributed equally to this work (Sérgio Almeida, Shagana T Balasubramaniam).

## Abstract

Pathogenic variants in O-GlcNAc transferase, which catalyses nucleocytoplasmic protein O-GlcNAcylation, cause a neurodevelopmental disorder (OGT-NDD) characterised by intellectual disability and motor impairments. However, mechanisms linking O-GlcNAc dyshomeostasis to brain development remain unclear. Longitudinal neuroimaging of an individual carrying the p.Asn648Tyr OGT-NDD variant, combined with developmental and molecular analyses of an *Ogt^N648Y/y^* mouse model, revealed that the mouse phenocopied the impaired cerebellar growth in the patient. Altered OGT distribution and reduced Calbindin expression preceded reduced Purkinje and granule cell densities and molecular layer thinning, alongside increased microglial density, persistent phagocytic cup-like structures and reduced interleukin (IL)-4 levels. Pharmacological OGT inhibition reduced IL-4-induced reporter activity *in vitro*, while primary *Ogt^N648Y/y^* microglia exhibited blunted IL-4-induced O-GlcNAcylation with enrichment of inflammatory and innate immunity programmes. These findings link impaired OGT catalytic function to postnatal cerebellar maturation during a critical developmental window and identify IL-4-associated microglial responses as a contributor to cerebellar pathology in OGT-NDD.

## Introduction

The cerebellum has long been recognized for its role in motor coordination and motor learning. However, a growing body of evidence reveals that it also contributes to cognition, language, affective regulation, and social behaviour through extensive reciprocal connections with cortical and subcortical regions^1–3^. Consequently, developmental disturbances affecting cerebellar structure and connectivity can have profound consequences beyond motor control. Indeed, cerebellar abnormalities are increasingly recognized as common features of neurodevelopmental disorders (NDDs), including autism spectrum disorder (ASD), attention-deficit/hyperactivity disorder (ADHD), and intellectual disability (ID), where cerebellar hypoplasia and vermian abnormalities frequently correlate with symptom severity^4,5^.

The cerebellum undergoes a prolonged and highly coordinated developmental programme that extends from embryogenesis into postnatal life. In humans, cerebellar development begins approximately 30 days post-conception and continues throughout the first years of life, whereas in mice many critical developmental events occur during the first three postnatal weeks and extend into postnatal days (P) 30–35^6,7^. During this period, the cerebellar cortex is organized into a highly structured laminar architecture comprising the external granule layer (EGL), molecular layer (ML), Purkinje cell layer (PCL), and internal granule layer (IGL). The EGL contains proliferating granule cell precursors (GCPs), which subsequently differentiate and migrate inward to populate the IGL as mature granule cells (GCs), the most abundant neuronal population in the brain. In parallel, Purkinje cells (PCs), whose somata form a monolayer within the PCL, undergo extensive dendritic growth into the ML, where they receive synaptic input from GC-derived parallel fibres, climbing fibers, and local interneurons. The mature cerebellar cortex is therefore organized into a stereotyped microcircuit in which excitatory parallel fibres from GCs provide input to PCs, the sole output neurons of the cerebellar cortex, enabling precise regulation of cerebellar function^8–10^. The prolonged postnatal period over which these developmental processes occur may render the cerebellum particularly vulnerable to genetic and environmental perturbations during early life.

Among cerebellar regions, the vermis has attracted particular attention in NDDs. It contributes to motor, cognitive, emotional, and social behaviours and is frequently affected in developmental conditions, including ASD, since reductions in vermal volume represent one of the most consistently reported cerebellar abnormalities^11^. Post-mortem studies of individuals with ASD have further revealed reduced numbers of PCs, highlighting the susceptibility of cerebellar circuits to developmental disruption^12,13^. Despite extensive characterisation of these anatomical abnormalities, the molecular and cellular mechanisms linking altered cerebellar development to circuit dysfunction associated with NDDs remain incompletely understood.

Beyond intrinsic neural developmental programmes, neuroimmune interactions play critical roles in shaping cerebellar maturation. Microglia, the most abundant immune cells of the central nervous system (CNS), contribute to neurogenesis, synaptic refinement, axonal remodelling, and clearance of apoptotic cells throughout development^14–17^. In the mouse cerebellum, the postnatal developmental window spanning from P10 to P30 overlaps with a period of heightened microglial activity, during which these cells participate in whole-cell phagocytosis^18,19^. While these processes are essential for normal brain development, dysregulated microglial activity can result in aberrant circuit remodelling underlying NDDs^20^.

Among the immune mediators regulating microglial function, interleukin (IL)-4, a type 2 cytokine classically associated with anti-inflammatory immune responses, is increasingly recognized for its functions in the CNS. IL-4 signalling promotes transcriptional programmes associated with tissue repair, neuronal survival, neurogenesis and regulation of microglial states^21–24^. In the postnatal cerebellum, IL-4 has been shown to restrain microglia-mediated neuronal engulfment, while elevated levels of this cytokine induce increased density of GCs and ADHD-like behaviours^18^.

A key regulator of cellular signalling during development is O-linked β-N-acetylglucosaminylation (O-GlcNAcylation), a dynamic and reversible post-translational modification involving the addition of a single N-acetylglucosamine moiety to serine and threonine residues of nucleocytoplasmic proteins^25^. O-GlcNAcylation regulates multiple cellular processes, including transcription, signal transduction, protein stability, stress responses, and synaptic plasticity^26,27^. Importantly, proteins involved in neural differentiation, cytoskeletal organization, synaptic maturation, and immune signalling are regulated by O-GlcNAcylation, highlighting its broad influence on CNS development^28–31^.

Cellular O-GlcNAcylation levels are maintained through the opposing activities of O-GlcNAc transferase (OGT), which catalyses addition of O-GlcNAc^32,33^, and O-GlcNAcase (OGA), which removes it^34,35^. Beyond its catalytic function, OGT also participates in protein scaffolding complexes and regulates proteolytic processing of host cell factor-1 (HCF-1), a transcriptional co-regulator involved in cell-cycle progression and developmental programmes^36^. Consistent with its fundamental biological function, complete deletion of *Ogt* results in embryonic lethality in mice^37^, highlighting the need for conditional approaches to investigate its role in CNS development.

A recent study has suggested a link between OGT and cerebellar development. In adult male mice, the cerebellum possesses the highest level of O-GlcNAcylation when compared to cortex and hippocampus, while conditional deletion of *Ogt* in GCPs induces abnormal cerebellar development and severe cerebellar hypoplasia^38^. Yet, the mechanisms underlying this phenotype remain unknown, and whether OGT supports cerebellar development through its catalytic activity, its non-catalytic functions, or both has not been determined.

Pathogenic variants in the *OGT* gene, residing on chromosome Xq13.1, cause a syndromic neurodevelopmental disorder termed OGT-NDD or O-GlcNAc Transferase Congenital Disorder of Glycosylation (OGT-CDG)^39–42^. Affected individuals frequently present global developmental delay, ID, epilepsy, ASD-related features, hypotonia, and motor impairments, highlighting the critical role of OGT in human brain development. Despite increasing recognition of the syndrome, the cellular and molecular mechanisms linking impaired O-GlcNAc homeostasis to circuit-level abnormalities remain poorly understood.

We recently described a *de novo* missense mutation in the catalytic domain of *OGT* (c. 1942A>T p.Asn648Tyr, *OGT^N648Y^*) in a male patient with OGT-NDD^43^. In addition to global developmental abnormalities, the patient exhibited clinical features consistent with impaired motor and cerebellar function, including truncal ataxia, persistent hypotonia, delayed motor development, absence of speech, sensory hypersensitivity, and behavioural dysregulation. We subsequently generated a mouse model carrying the corresponding *Ogt*^N648Y/y^ variant, which displays key features of the patient symptoms, including reduced brain size, disrupted O-GlcNAc homeostasis, and sensory-motor deficits, including impaired performance in tasks requiring fine motor coordination^44^. Together, these findings suggest that disruption of OGT catalytic function may particularly affect cerebellar developmental programmes.

Here, we investigated the effects of the OGT*^N648Y^* variant for postnatal cerebellar development and neuroimmune regulation. Longitudinal neuroimaging of the patient carrying the OGT^N648Y^ variant revealed striking cerebellar hypoplasia, with pronounced involvement of the cerebellar vermis and impaired postnatal cerebellar growth. This phenotype was recapitulated in the *Ogt*^N648Y/y^ mouse model, in which cerebellar hypoplasia emerged during postnatal maturation and was accompanied by age- and layer-dependent alterations in OGT abundance and progressive disruption of O-GlcNAc homeostasis. Early changes at P10 included reduced calbindin signal intensity and altered OGT distribution across the cerebellar cortex, preceding the reduction in PC and GC densities and ML thickness observed at P30. These neuronal alterations occurred alongside increased microglial abundance, persistence of phagocytic cup-like structures and altered microglia–PC interactions. At P30, mutant cerebella additionally showed reduced IL-4 levels and inflammatory pathway enrichment in cerebellar microglia. We further found that OGT activity contributes to IL-4-associated signalling in microglia, with OGT inhibition reducing IL-4-dependent signalling and *Ogt*^N648Y/y^ primary microglia showing a blunted O-GlcNAc response to IL-4 stimulation. Together, these findings indicate that impaired OGT catalytic function becomes increasingly consequential during postnatal cerebellar maturation, coinciding with progressive disruption of O-GlcNAc homeostasis and altered neuroimmune regulation, and implicate defective IL-4-associated microglial responses as a contributor to cerebellar pathology in OGT-NDD.

## Results

### Postnatal cerebellar hypoplasia and disrupted O-GlcNAc homeostasis in Ogt^N648Y/y^ mice

A previously described patient harbouring the *OGT* p.Asn648Tyr variant within the catalytic domain ^39^ underwent longitudinal magnetic resonance imaging (MRI) at three time points: twice between 0–4 years of age and once between 13–18 years of age. MRI analysis revealed persistent abnormalities of the posterior fossa, including mega cisterna magna, reduced cerebellar vermis size, and mild pontocerebellar atrophy (**Fig. 1A**). These abnormalities were already evident in early childhood and showed mild progression over time, with gradual reduction in pontine and cerebellar volume, widening of cerebellar sulci, and persistent vermian hypoplasia. Linear measurements further supported impaired cerebellar growth, with cerebellar length remaining unchanged at 22 mm across all three time points, while pontine length increased from 17 to 17.5 and 18 mm and cerebral anteroposterior length from 148 to 151 and 155 mm, respectively. **(Fig. 1A).** Mild cortical and subcortical volume loss was also noted, particularly in the occipital regions. At the latest imaging time point, pontocerebellar atrophy remained mild but progressive, accompanied by a shortened corpus callosum and occipital bone thickening.

**Figure 1.**
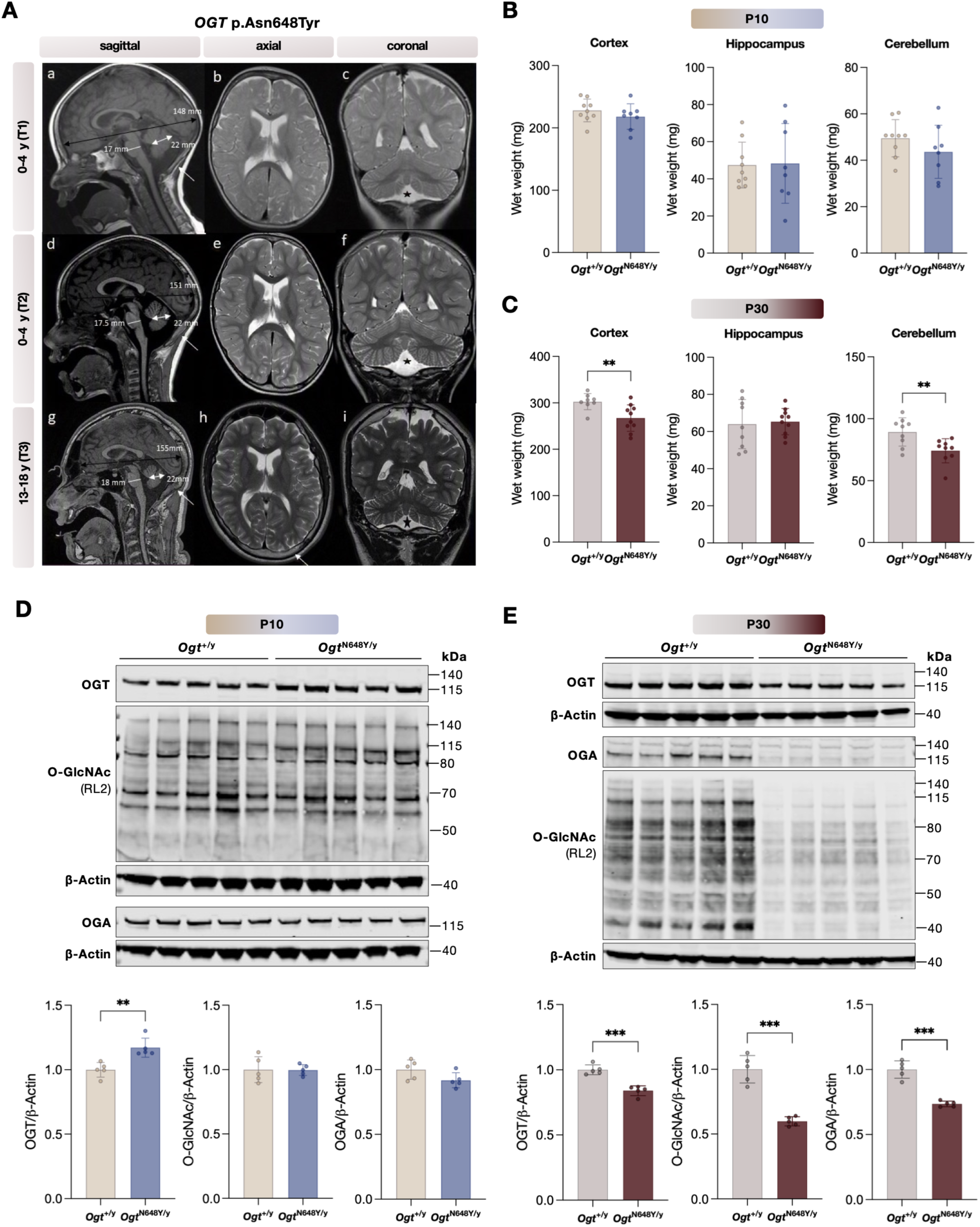
OGT^N648Y^ mutation causes cerebellar hypoplasia in OGT-NDD patient and impairs postnatal cerebellar growth in mice. (A) Serial brain MRI scans from an OGT-NDD patient harbouring the *OGT* p.Asn648Tyr variant at 0–4 years (T1; a–c), 0–4 years (T2; d–f), and 13–18 years (T3; g–i) demonstrate thickened occipital bones (white arrows), pontocerebellar atrophy (black arrows), thinning of the corpus callosum, hypoplasia of the cerebellar vermis, and enlargement of the cisterna magna (asterisk). (B-C) Brain region wet weights in WT and Ogt^N648Y/y^ mice at P10 and P30. Cortex, hippocampus, and cerebellum were dissected and weighed at postnatal day 10 (P10) and postnatal day 30 (P30). (D-E) Western blot analysis of O-GlcNAc, OGT, and OGA levels (RL2) in cerebellum at P10 (D) and P30 (E) (n = 5). All data are represented as mean ± SD and each data point represents a mouse. Unpaired t-test with Welch’s correction is used for statistics. Significance is shown as: *p < 0.05, **p < 0.01 and ***p < 0.001.

To assess whether the *Ogt*^N648Y/y^ mouse model recapitulates the abnormalities observed in the patient, cortex, hippocampus, and cerebellum were dissected at P10 and P30, representing early and late stages of cerebellar development (**Fig. 1B-1C**). While absolute brain region wet weights were unaffected at P10, *Ogt*^N648Y/y^ mice exhibited reduced cortical and cerebellar mass compared to wild-type (WT) littermate controls at P30, indicating an age-dependent emergence of a neurodevelopmental phenotype in the *Ogt*^N648Y/y^ mouse model, recapitulating key neuroanatomical features observed in the patient.

Since the *OGT* p.Asn648Tyr variant has been shown to compromise OGT catalytic activity *in vitro* ^41^, we next examined whether the onset of cerebellar hypoplasia was associated with altered O-GlcNAcylation in the *Ogt*^N648Y/y^ mice. OGT, OGA, and global O-GlcNAc levels were assessed by immunoblotting in cerebellar lysates at P10 and P30 (**Fig. 1D-1E**). At P10, O-GlcNAc levels were not significantly altered in *Ogt*^N648Y/y^ cerebella, despite an increased OGT, suggesting that O-GlcNAc homeostasis is largely maintained at this early developmental stage. Notably, this increase in cerebellar OGT differs from our previous observations in whole-brain lysates from *Ogt*^N648Y/y^ mice^44^, indicating context-dependent regulation of OGT expression across tissues and developmental stages. In contrast, at P30, mutant cerebella showed a significant reduction in global O-GlcNAc levels compared to WT controls. This reduction was accompanied by decreased levels of both OGT and OGA, indicating disruption of the O-GlcNAc cycling machinery during postnatal cerebellar maturation.

Together, these findings show that the *Ogt^N648Y/y^* mouse model recapitulates the cerebellar phenotypes observed in the OGT^N648Y^, associated with an age-dependent reduction in cerebellar mass that coincides with impaired maintenance of O-GlcNAc homeostasis.

### Ogt^N648Y/y^ mice exhibit altered postnatal cerebellar maturation

We hypothesized that the reduction in cerebellar wet weight observed in *Ogt*^N648Y/y^ mice at P30 could reflect impaired postnatal growth. To determine whether cerebellar cytoarchitecture was affected, and given the persistent vermian hypoplasia observed in the patient, we focused on the cerebellar vermis, specifically lobules IV/V, which receive projections from motor areas of the cerebral cortex and are involved in motor coordination^45^. Sagittal cerebellar sections from WT and *Ogt*^N648Y/y^ mice at P10 and P30 were immunostained for Pax6 to identify granule lineage cells and calbindin to identify PCs and delineate the ML (**Fig. 2A-B**).

**Figure 2.**
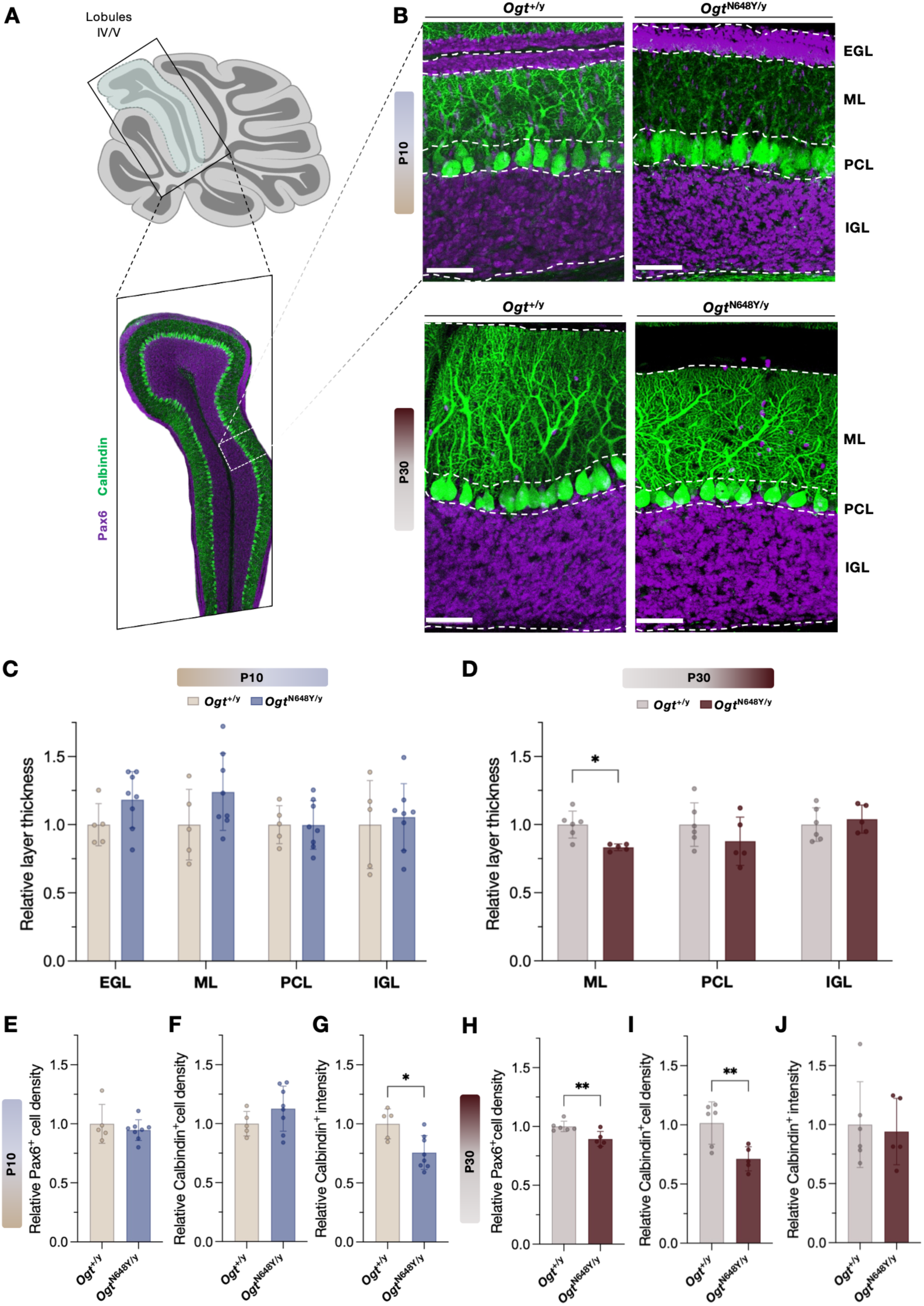
*Ogt*^N648/Y^ mice exhibit aberrant cerebellar postnatal development. (A) Schematic overview of the analysed cerebellar region. Cerebellar vermis lobules IV/V were selected for histological assessment. The representative confocal image shows Calbindin^+^ Purkinje cells and dendrites outlining the Purkinje cell and molecular layers (green) and Pax6^+^ granule cells in the granule cell layer (purple). (B) Representative images of slices of the cerebellar vermis of lobule IV/V stained with anti-Calbindin (Purkinje cells) and anti-Pax6 (granule cells) at P10 and P30. (C-D) Quantification of relative thickness of cerebellar cortex layers normalized to WT controls. Quantitative analysis of layer thickness normalised to Ogt+/y mice. The thickness of each layer was measured in the area outlined with a dashed box in (B) lobule 4/5. (E-F) Quantification of the relative number of Calbindin^+^ cells (Purkinje cells) and Pax6^+^cells (granule cells) per µm2 at P10 normalized to WT controls. (G-H) Quantification of the relative number of Calbindin^+^ cells (Purkinje cells) and Pax6^+^cells (granule cells) per µm2 at P30 normalized to WT controls. EGL = external granular layer, ML = molecular layer, PCL = Purkinje cell layer, IGL = internal granule cell layer. Data are shown as mean ± SD. Two-way ANOVA (C-D) and unpaired t-test with Welch’s correction (E-H) is used for statistics: *p < 0.05, **p < 0.01, ***p < 0.001. Scale bars, 50 µm. Each data point represents one mouse; values correspond to the mean quantification from three sections per mouse.

We first quantified the thickness of the major cerebellar cortical layers. At P10, when GC precursors are still proliferating within the EGL and migrating inward to populate the IGL, no significant differences in layer thickness were detected between WT and *Ogt*^N648Y/y^ mice (**Fig. 2C**). By contrast, at P30, when cerebellar lamination is largely established and the EGL has disappeared, mutant mice displayed a significant reduction in ML thickness, whereas the PCL and IGL were not significantly altered (**Fig. 2D**), indicating that overt laminar abnormalities emerge during postnatal cerebellar maturation.

We next assessed whether these architectural changes were accompanied by alterations in the major neuronal populations. At P10, granule lineage cell and PC densities were comparable between genotypes (**Fig. 2E-F**). However, calbindin fluorescence intensity in the ML was significantly reduced in *Ogt*^N648Y/y^ cerebella at this stage (**Fig. 2G**), suggesting an early alteration in the PC compartment despite preserved PC density. At P30, mutant mice showed a significant reduction in both PC and GC density compared with WT controls (**Fig. 2H-J**). Together, these findings indicate that changes in PC-associated calbindin signal are already detectable at P10 and precede the reduction in neuronal densities and ML thickness observed at P30.

Given the increased OGT expression detected in cerebellar lysates at P10, we next examined whether the *Ogt*^N648Y/y^ variant altered the layer-specific distribution of OGT in the cerebellum (**Supplementary Fig. 1A**). Across both genotypes, the highest mean OGT fluorescence intensity was detected in the PCL, consistent with prominent OGT-positive cells surrounding the Calbindin-positive PCs (**Supplementary Fig. 1B**). Comparison between genotypes revealed a selective increase in OGT intensity within the EGL of *Ogt*^N648Y/y^ mice, whereas OGT intensity in the ML, PCL, and IGL remained largely comparable with WT controls (**Supplementary Fig. 1B-C**). Finally, we quantified the area-weighted contribution of each layer to the total OGT signal within the analysed region-of-interest (ROI). Although the IGL accounted for the largest proportion of total OGT in both genotypes, its relative contribution was significantly reduced from 41% to 35% in *Ogt*^N648Y/y^ mice (**Supplementary Fig. 1D**). Conversely, the relative contribution of the EGL to the total OGT signal was significantly increased in *Ogt*^N648Y/y^ mice (**Supplementary Fig. 1E**). These findings indicate that the *Ogt*^N648Y/y^ mutation shifts OGT distribution towards increased EGL-associated and reduced IGL-associated OGT signal.

Together, these data show that *Ogt*^N648Y/y^ cerebella at P10 already exhibit early molecular alterations, including reduced PC-associated calbindin signal and layer-specific redistribution of OGT, despite preserved gross cytoarchitecture. By P30, these early changes are accompanied by reduced ML thickness and decreased PC and GC densities, consistent with an age-dependent disruption of postnatal cerebellar maturation.

### Ogt^N648Y/y^ mice exhibit a late postnatal presence of microglial phagocytic structures and impaired IL-4 signalling

The reduction in PC and GC densities at P30 suggested that dysregulated postnatal cerebellar maturation in *Ogt*^N648Y/y^ mice may involve altered cell elimination and/or circuit refinement. To determine whether these changes were associated with altered microglial abundance and phagocytic structures, we quantified IBA1-positive microglia in vermal lobules IV/V of WT and *Ogt*^N648Y/y^ mice at P10 and P30. Phagocytic cups were defined as cup-like IBA1-positive structures located at the distal ends of microglial processes, consistent with structures previously associated with cellular engulfment^18,19,46^ (**Fig. 3A**).

**Figure 3:**
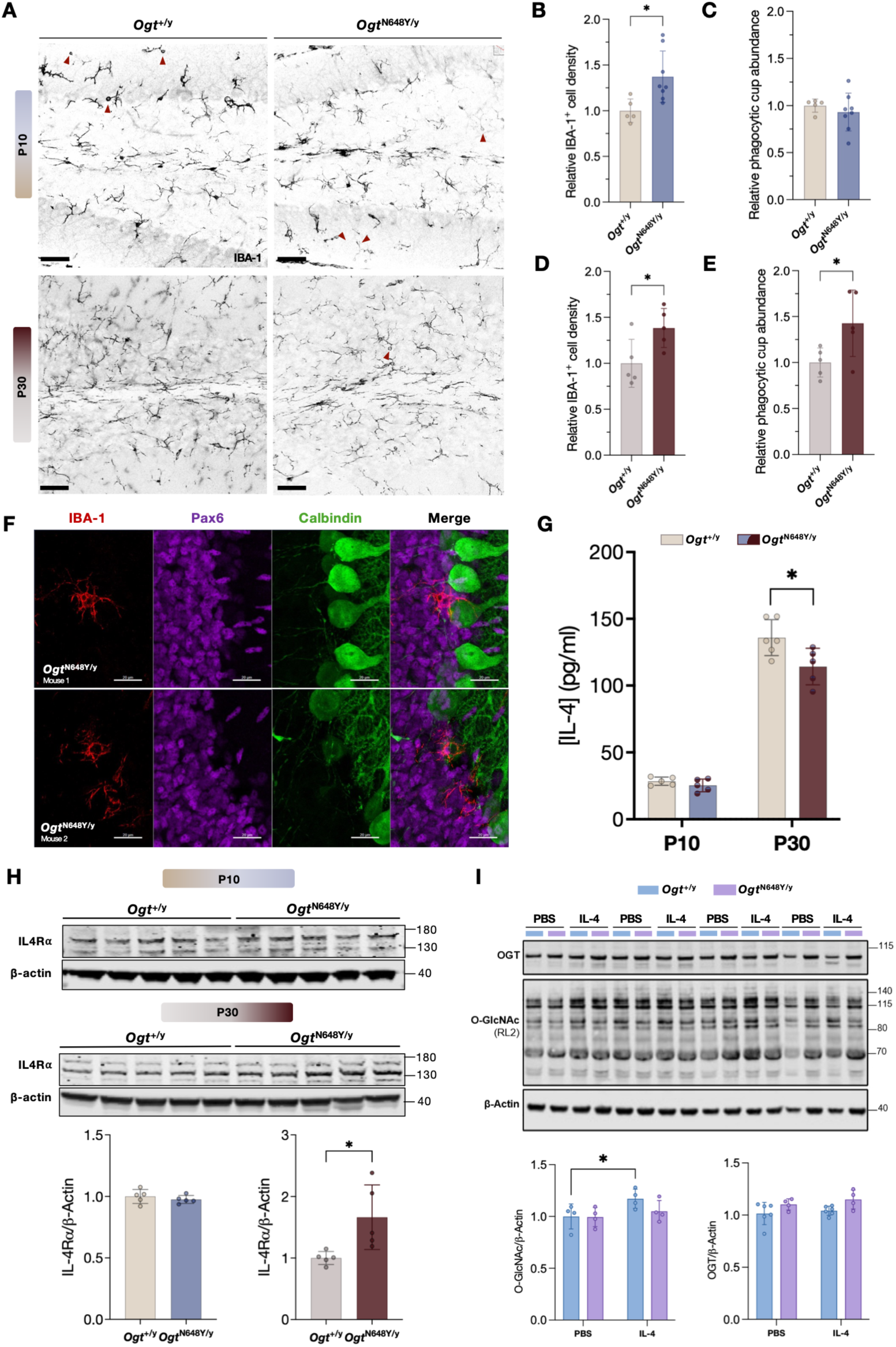
Altered IL-4 signalling at P30 correlates with persistent microglial phagocytosis in *Ogt*^N648Y/y^ cerebellum. (A) Representative images of slices of the cerebellar vermis of lobules IV/V stained with anti-IBA-1 (microglia) at P10 and P30. Red arrowheads indicate the presence of phagocytic cups. Scale bar: 50 µm (B-C) Quantification of number of IBA-1^+^ cells (microglia) and phagocytic cups per µm^2^ of lobules IV/V normalized to WT controls at P10. (D-E) Quantification of number of IBA-1^+^ cells (microglia) and phagocytic cups per µm^2^ of lobules IV/V normalized to WT controls at P30. (F) Representative images of IBA-1^+^ microglia in contact with and forming phagocytic cups around Calbindin^+^ Purkinje cells in *Ogt*^N648/Y^ mice at P10. Red: IBA-1, Purple: Pax6, Green: Calbindin. (G) IL-4 levels measured by ELISA in cerebellar lysates at P10 and P30. (H) Western blot analysis of IL-4 receptor alpha (IL-4Rα) expression in cerebellar lysates at P10 and P30. (I) Western blot analysis and quantification of OGT and global O-GlcNAcylation (RL2) in primary microglia isolated from P5 whole brain WT (n=4) and *Ogt*^N648Y/y^ (n=4) mice and stimulated with PBS or IL-4 for 8 h. Relative levels were normalized to β-actin. Molecular weights are indicated in kDa. Data are shown as mean ± SD. Unpaired t-test (A-E and H) and multiple unpaired t-test with Welch’s correction (G and I) and is used for statistics: *p < 0.05, **p < 0.01, ***p < 0.001.

At P10, *Ogt*^N648Y/y^ mice displayed a significant increase in IBA1-positive microglial density compared with WT littermate controls (**Fig. 3B**). However, the abundance of phagocytic cup-like structures normalised to the analysed area was not significantly different between genotypes at this stage (**Fig. 3C**), indicating that the early increase in microglial density was not accompanied by a global increase in phagocytic cups. Notably, IBA1-positive microglial processes and cup-like structures were observed in close proximity to calbindin-positive PCs in *Ogt*^N648Y/y^ mice at P10, whereas comparable interactions were not detected in WT mice (**Fig. 3F)**. Such microglia–PC contacts may reflect altered surveillance or interaction with PCs during postnatal cerebellar maturation, although static imaging does not allow confirmation of active engulfment.

At P30, *Ogt*^N648Y/y^ mice continued to show increased IBA1-positive microglial density in the cerebellar vermis (**Fig. 3D**). In contrast to P10, this was accompanied by a significant increase in the number of phagocytic cups (**Fig. 3E**), rarely present in microglia at this age^18^, indicating that *Ogt*^N648Y/y^ microglia retain or acquire elevated phagocytic structures at a developmental stage by which cerebellar maturation is largely established. Together with the reduced ML thickness and decreased PC and GC densities observed at P30, these findings indicate that altered microglial dynamics emerge at the same developmental stage at which impaired cerebellar maturation becomes apparent in *Ogt*^N648Y/y^ mice.

We next sought to identify a potential underlying mechanism contributing to the increase in microglial phagocytic structures observed in *Ogt*^N648Y/y^ mice at P30. IL-4 signalling represented a particularly relevant candidate pathway, as IL-4 has recently been shown to regulate microglia-dependent whole-cell engulfment during postnatal cerebellar development. Low IL-4 coincide with periods of high microglial engulfment, whereas rising IL-4 levels contribute to restricting further pruning as cerebellar circuits mature^18^. We therefore measured IL-4 levels in cerebellar lysates from WT and *Ogt*^N648Y/y^ mice at P10 and P30 by ELISA to determine whether the microglial phenotype was associated with altered IL-4 signalling. Consistent with the developmental increase in IL-4 during cerebellar maturation, IL-4 levels increased markedly from P10 to P30 (**Fig. 3G**). At P10, IL-4 levels were comparable between the genotypes, whereas at P30 they were significantly reduced in *Ogt*^N648Y/y^ cerebella compared with WT littermate controls (**Fig. 3G**). This decrease was not observed in either the cortex or hippocampus (**Supplementary Fig. 2A**), suggesting that the altered IL-4 abundance is specific to the cerebellum. The reduction in cerebellar IL-4 coincided with the stage at which *Ogt*^N648Y/y^ mice displayed increased phagocytic cup abundance, suggesting that insufficient IL-4 availability could contribute to the persistent presence of microglial phagocytic cup-like structures in the *Ogt*^N648Y/y^ cerebellum.

We next assessed whether reduced IL-4 levels were accompanied by altered expression of IL4Rα, the essential subunit of the IL-4 receptor. Immunoblot analysis of cerebellar lysates showed no significant difference in IL4Rα levels between genotypes at P10 (**Fig. 3H**). By contrast, IL4Rα protein abundance was increased in *Ogt*^N648Y/y^ cerebella at P30 (**Fig. 3H**), potentially reflecting a compensatory response to reduced IL-4 availability. However, despite elevated IL4Rα expression, *Ogt*^N648Y/y^ mice continued to exhibit increased microglial phagocytic cup abundance at P30, indicating that receptor upregulation may not be sufficient to normalise microglial phagocytic activity.

To examine whether altered IL-4-associated signalling was accompanied by changes in O-GlcNAc homeostasis, we analysed primary microglia isolated from P5 WT and *Ogt*^N648Y/y^ mice after one week in culture and stimulated with IL-4 for 8 h. In WT primary microglia, IL-4 stimulation increased global O-GlcNAcylation, whereas this response was blunted in *Ogt*^N648Y/y^ cultures, despite no marked differences in OGT protein abundance (**Fig. 3I**). Thus, *Ogt*^N648Y/y^ microglia fail to elicit a normal O-GlcNAc response to IL-4 stimulation in culture.

Additional *in vitro* experiments further supported impaired IL-4 signalling output upon disruption of OGT activity. Pharmacological inhibition of OGT with OSMI-4 reduced IL-4-induced STAT6-dependent SEAP activity in HEK-Blue IL-4/IL-13 reporter cells^47,48^, whereas inhibition of OGA with Thiamet-G did not significantly alter the reporter response (**Supplementary Fig. 2B**). These findings indicate that OGT activity contributes to efficient IL-4 signalling output, while increasing global O-GlcNAcylation through OGA inhibition is not sufficient to modify the response. Likewise, after 24 h of IL-4 stimulation, primary *Ogt*^N648Y/y^ microglia showed a blunted increase in global O-GlcNAcylation and reduced induction of the IL-4-responsive marker ARG1^49^ compared to WT cells (**Supplementary Fig. 2C**) Together, these findings indicate that OGT activity and an appropriate O-GlcNAc response are required for efficient IL-4-associated signalling, whereas increasing global O-GlcNAcylation by OGA inhibition alone is insufficient to enhance pathway output. This impaired OGT-dependent response may contribute to the persistence of microglial phagocytic cup-like structures observed in *Ogt*^N648Y/y^ cerebella.

Collectively, these findings show that the increased microglial phagocytic phenotype in *Ogt*^N648Y/y^ cerebella is accompanied by reduced IL-4 availability, increased IL4Rα abundance, and an impaired OGT-dependent response to IL-4 stimulation. The increase in IL4Rα despite impaired IL-4 responsiveness suggests that the signalling defect lies downstream of receptor availability and points to a requirement for OGT in efficient propagation of the IL-4 response. Collectively, these findings suggest that disrupted IL-4–OGT signalling may contribute to the persistence of microglial phagocytic activity during postnatal cerebellar maturation.

### Ogt^N648Y/y^ cerebellar microglia display an inflammatory transcriptional profile

The increased microglial density, persistent presence of phagocytic structures, and altered IL-4 signalling suggest that *Ogt*^N648Y/y^ microglia may acquire an altered functional state during postnatal maturation. To investigate this, we performed bulk RNA sequencing of microglia from P30 WT and *Ogt*^N648Y/y^ cerebella.

Principal component analysis (PCA) of the 2,000 most variable genes revealed partial segregation of WT and *Ogt*^N648Y/y^ samples, consistent with genotype-associated differences in the transcriptomic profile of isolated cerebellar microglia (**Fig. 4A**). Similarly, hierarchical clustering of genes ranked by nominal p-value largely separated samples according to genotype (**Fig. 4C**).

**Figure 4.**
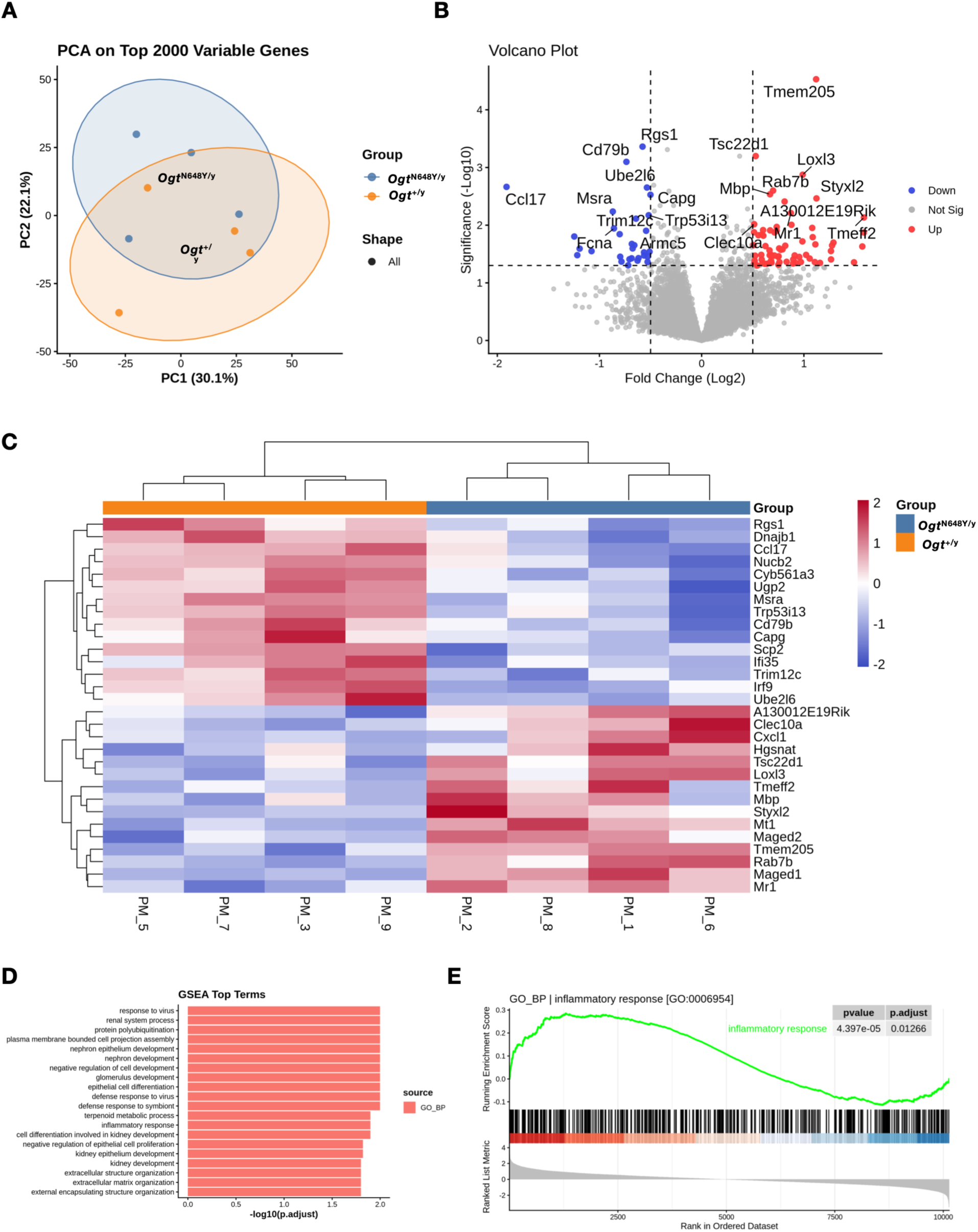
*Ogt*^N648Y/y^ induces a pro-inflammatory transcriptional profile in cerebellar microglia. (A) Principal component analysis (PCA) of the 2,000 most variable genes from bulk RNA sequencing of cerebellar microglia isolated from P30 WT and mutant mice. (B) Volcano plot showing differential gene expression between WT and *Ogt^N648Y/y^* microglia. Red and blue points indicate genes displaying nominally increased or decreased expression, respectively, whereas grey points indicate non-significant genes. No genes significantly differentially expressed following multiple testing correction. (C) Heatmap of the top 30 genes ranked by nominal p-value. Unsupervised hierarchical clustering largely separates WT and *Ogt*^N648Y/y^ microglia according to genotype. (D) Gene set enrichment analysis (GSEA) of Gene Ontology Biological Process (GO_BP) terms using the ranked transcriptomic dataset. (E) Enrichment plot for the inflammatory response gene set (GO:0006954), demonstrating significant enrichment in *Ogt*^N648Y/y^ microglia (adjusted p = 0.0169). Data were generated from microglia isolated from the cerebella of P30 WT (n = 4) and *Ogt*^N648Y/y^ (n = 4) mice.

Differential expression analysis identified several genes with nominal expression changes between WT and *Ogt*^N648Y/y^ microglia, however, no individual genes remained significantly different after correction for multiple testing (**Fig. 4B**). We therefore used gene set enrichment analysis (GSEA) to assess coordinated pathway-level changes across the ranked transcriptome. GSEA revealed significant enrichment of biological processes associated with inflammatory and innate immune responses in *Ogt*^N648Y/y^ microglia, including inflammatory response, response to virus, defence response to virus, and defence response to symbiont (**Fig. 4D**). Additional enriched pathways included protein polyubiquitination and extracellular structure organisation, indicating broader alterations in protein regulatory pathways and cellular remodelling.

Notably, the inflammatory response gene set (GO:0006954) was significantly enriched in *Ogt*^N648Y/y^ microglia (adjusted p = 0.01266; **Fig. 4E**). Genes contributing to this enrichment included cytokines and chemokines such as *Il1a, Il1b, Tnf, Ccl2, Ccl4, Ccl7, Cxcl1, Cxcl2,* and *Cxcl10*; innate immune and inflammatory regulators *Tlr2, Cd14, Nlrp3, Nfkbia, Nfkbiz, Nfkb2, Rel, Socs3,* and *Zfp36*; complement-associated genes *C3, C3ar1, C5ar1,* and *C5ar2*; and microglial activation- and phagocytosis-associated genes including *Apoe, Tyrobp, Fcer1g, Clec7a, Axl, Cst7,* and *Lgals1* (**Supplementary Table 1**).

Together, these data indicate that *Ogt*^N648Y/y^ cerebellar microglia display coordinated enrichment of inflammatory transcriptional programmes at P30, consistent with the increased microglial abundance, altered phagocytic morphology, and impaired IL-4-associated regulation observed in the *Ogt*^N648Y/y^ cerebellum.

## Discussion

In this study, we link the pathogenic OGT^N648Y^ variant to impaired postnatal cerebellar maturation and identify altered neuroimmune regulation as one component of this phenotype. Longitudinal MRI of the affected individual revealed persistent vermian hypoplasia and mild progressive pontocerebellar atrophy, while the corresponding *Ogt*^N648Y/y^ mouse model developed reduced cerebellar mass during postnatal maturation. This developmental phenotype was accompanied by disruption of O-GlcNAc homeostasis, altered cerebellar cortical organisation, reduced PC and GC densities, increased microglial abundance and phagocytic structures, reduced cerebellar IL-4 levels, impaired IL-4-associated O-GlcNAc responses, and inflammatory pathway enrichment in isolated cerebellar microglia. Together, these findings suggest that the OGT^N648Y^ mutation affects cerebellar development through a combination of cell-intrinsic alterations and disrupted neuroimmune interactions.

The cerebellum may be particularly susceptible to impaired OGT function as its postnatal development requires extensive proliferation, differentiation and circuit remodelling during a period of high O-GlcNAc activity. In the affected individual, cerebellar length remained unchanged at 22 mm across the longitudinal imaging period, despite continued growth of the pons and cerebrum, supporting sustained impaired cerebellar growth rather than global developmental delay. A similar developmental trajectory was observed in the *Ogt*^N648Y/y^ mice, where cerebellar mass was preserved at P10 but reduced by P30, placing the emergence of the phenotype within a developmental window broadly corresponding to the prolonged period of cerebellar maturation extending through infancy and early childhood in humans.

Previous studies provide a mechanistic basis for this vulnerability. OGT and O-GlcNAcylation increase throughout the early postnatal cerebellar cortex, with particularly high O-GlcNAc levels in proliferating GCPs in the EGL, and deletion of *Ogt* from GCPs disrupts cerebellar growth through impaired SHH-GLI2 signalling^50^. Moreover, the cerebellum exhibits particularly high levels of O-GlcNAcylation compared with other brain regions, and loss of OGT in the granule cell lineage causes reduced cerebellar size, abnormal GC distribution, disrupted PC and Bergmann glial organisation and altered synaptic connectivity through Gα12–RhoA/ROCK signalling^38^. These genetic loss-of-function models demonstrate an essential requirement for OGT during cerebellar development but rely on depletion of OGT from defined cell populations. In contrast, *Ogt*^N648Y/y^ mice retain OGT expression despite impaired catalytic function, demonstrating that partial disruption of OGT activity is sufficient to perturb cerebellar development. Our findings therefore extend previous genetic studies beyond OGT depletion, and suggest that, in a model harbouring a clinical OGT variant, the prolonged postnatal growth of the cerebellum together with its dependence on O-GlcNAc-regulated developmental pathways render it as particularly vulnerable to partial disruption of OGT catalytic activity.

Our data further suggest that OGT dysregulation begins before overt cerebellar hypoplasia becomes evident. Although gross lamination and neuronal densities were preserved at P10, calbindin intensity was already reduced in mutant cerebella, indicating an early alteration of PCs. Notably, global O-GlcNAc levels remained preserved at this stage, whereas OGT protein abundance was already increased, suggesting that O-GlcNAc homeostasis is maintained under increased compensatory pressure before structural abnormalities emerge.

At the same age, OGT distribution was markedly heterogeneous across the cerebellar layers. In *Ogt*^N648Y/y^ cerebella, OGT intensity was selectively increased in the EGL, while levels in the ML, PCL and IGL remained comparable with WT controls. When the contribution of each layer to the total OGT signal was considered, the relative contribution of the EGL was increased, whereas that of the IGL was reduced. Thus, the early increase in cerebellar OGT appears to reflect a shift towards greater EGL-associated OGT rather than broadly elevated OGT across the cerebellar cortex.

Across both genotypes, the strongest OGT signal was detected in the PCL, including prominent OGT-positive signal in cells surrounding PC somata. Given the close association of Bergmann glial cell bodies with PCs within this layer, this local OGT signal reflects contributions from both neuronal and glial populations. A direct contribution of OGT dysfunction to PC development should also be considered, as PC-specific loss of OGT causes dendritic and spine degeneration during postnatal maturation and ultimately PC loss, associated with mitochondrial dysfunction and increased ROS generation^51^. Together with the reduced calbindin signal observed at P10, which suggests calcium dysfunction in PCs^52^, these findings propose that these neurons, and potentially their associated glial environment, are affected before detectable changes in PC number emerge. The subsequent reduction in both PC and GC densities is therefore likely to reflect interacting neuronal, glial and neuroimmune mechanisms rather than a solely microglia-mediated process.

The timing of the microglial phenotype may further support a model in which microglia respond to an underlying defect in PC maturation. Physiological microglial engulfment of PCs is most prominent during the early postnatal period, around P3–P4 ^53^. However, *Ogt*^N648Y^ mice display preserved PC density at P10, followed by reduced PC density at P30. Together with increased microglial density at P10 and enhancement of phagocytic structures at P30, these results raise the possibility that developmentally compromised PCs persist beyond the normal period of PC elimination and remain subject to delayed or prolonged microglial engagement. The close association of IBA-1-positive processes and cup-like structures with PCs at P10 should not be interpreted as direct evidence of PC engulfment. Cerebellar microglia can dynamically survey PC somata and dendritic arbours *in vivo*, establishing contacts that can persist for prolonged periods^46^. Microglia also influence developmental refinement of PC circuitry through mechanisms independent of direct engulfment; for example, microglial depletion impairs climbing fibre elimination by reducing GABAergic inhibition onto PCs rather than through loss of direct microglial phagocytosis of climbing fibres^47^. Thus, early microglia-PC interactions may reflect altered surveillance or regulation of dysfunctional low-calbindin PCs, whereas the later increase in phagocytic structures is consistent with a broader clearance-associated phenotype. This distinction is important because microglia can influence cerebellar maturation through direct neuronal contact, cytokine-mediated signalling, regulation of synaptic refinement and phagocytosis^18,53–55^. Together, these observations support a model in which altered PC maturation is accompanied by prolonged microglial engagement during postnatal cerebellar development.

The developmental regulation of IL-4 provides a potential link between these altered microglial features and OGT dysfunction. It has previously been demonstrated that cerebellar IL-4 concentrations increase during postnatal maturation as microglial engulfment of neuronal material declines, and that increasing IL-4 during the early postnatal period suppresses microglia-dependent GC pruning. Importantly, this effect requires IL4Rα signalling in microglia, as the effects of elevated IL-4 on cerebellar pruning were abolished in *Il4rα*^fl/fl^::*Cx3cr1*-CreER mice^18^. In our study, the normal developmental increase in cerebellar IL-4 was attenuated in *Ogt*^N648Y/y^ mice at P30, coinciding with increased microglial phagocytic structures and reduced neuronal densities. The accompanying increase in IL4Rα protein may therefore reflect compensatory responsiveness to reduced ligand availability. However, IL4Rα is not restricted to microglia. Neuronal IL4Rα regulates synaptic transmission and neuronal homeostasis in other CNS regions, and recent mapping of the IL-4 receptor system has also identified IL4Rα expression in cerebellar PCs^56,57^. Consequently, the increased IL4Rα expression observed in whole-cerebellar lysates may reflect responses occurring in several cerebellar cell populations and cannot be attributed exclusively to microglia.

An additional question raised by the reduced IL-4 concentration is the cellular origin of IL-4 in the developing cerebellum, which remains incompletely defined. In other contexts, IL-4 can be produced by meningeal CD4^+^ T cells, while neuronal IL-4 expression has also been demonstrated, particularly under conditions of neuronal stress or neuroinflammation^21,58^. This issue may be particularly relevant to OGT-NDD because the OGT^N648Y^ mutation is present in all cell types and could therefore affect both IL-4-responsive cells and those responsible for producing the cytokine. In type 2 immune responses, IL-4 signalling through IL4Rα–STAT6 promotes GATA3-dependent programmes that reinforce IL-4 production, creating a positive regulatory loop^59,60^. Disruption of OGT-dependent signalling in an IL-4-producing population could therefore potentially contribute to the reduced IL-4 availability observed in mutant cerebella, in addition to the impaired response of microglia to the cytokine. Distinguishing cerebellum-intrinsic effects from systemic consequences of the mutation will ultimately require region- or cell-type-restricted models of *Ogt*^N648Y/y^. Such approaches will also be important for defining the cellular origin and regulation of IL-4 during cerebellar maturation.

Our cellular experiments provide further evidence that OGT activity is required for an appropriate IL-4 response. *In vitro*, pharmacological inhibition of OGT with OSMI-4 reduced IL-4-induced reporter activity, whereas inhibition of OGA with Thiamet-G did not significantly alter the response. This distinction suggests that IL-4 signalling depends on OGT activity or dynamic, substrate-specific O-GlcNAcylation rather than simply on increasing global O-GlcNAc levels. Consistent with this interpretation, IL-4 stimulation increased global O-GlcNAcylation in WT primary microglia after 8 h, whereas this response was absent in *Ogt*^N648Y/y^ cells. At 24 h, mutant microglia also showed reduced induction of ARG1, indicating that the impaired O-GlcNAc response is accompanied by reduced downstream IL-4-responsive transcription. Previous studies similarly showed that OGT inhibition attenuates IL-4/IL-13-associated macrophage programmes and that STAT6 itself can be directly O-GlcNAcylated^61,62^. Notably, O-GlcNAcylation of the STAT6 transactivation domain modulates its transcriptional activity without preventing IL-4-induced Y641 phosphorylation^62^. This provides a potential explanation for how OGT dysfunction could reduce IL-4 signalling output without necessarily disrupting the initial phosphorylation-dependent activation of STAT6. Overall, our results support a model in which the *Ogt*^N648Y/y^ mutation does not abolish IL-4 signalling but reduces the efficiency with which IL-4 stimulation is translated into an appropriate O-GlcNAc-dependent cellular response.

This impaired response may be particularly relevant to the developmental timing of the microglial phenotype. Under normal conditions, increasing IL-4 accompanies the transition away from the highly phagocytic early postnatal state of cerebellar microglia. In *Ogt*^N648Y/y^ mice, both components of this regulatory system appear altered at P30, where IL-4 availability is reduced and microglia show an impaired OGT-dependent response to IL-4. These changes may reduce the effectiveness of the signal that normally accompanies termination of developmental microglial remodelling, providing a plausible framework for the persistence of phagocytic structures at P30 while remaining compatible with additional roles of microglia in neuronal surveillance and circuit regulation. Interestingly, the transcriptomic profile of isolated P30 cerebellar microglia is consistent with this altered functional state. Although no individual genes remained significant after correction for multiple testing, GSEA identified coordinated enrichment of inflammatory and innate immune response programmes. The genes contributing to this signature encompassed cytokine and chemokine signalling, innate immune receptors, complement-associated pathways and microglial activation-related machinery, indicating a broad shift in immune state. The presence of complement-associated genes is of interest given the established role of complement signalling in microglia-mediated synaptic pruning and circuit refinement in other regions of the developing CNS^53,63,64^. Overall, the pathway-level changes are consistent with the increased microglial abundance and altered microglial morphology observed histologically and suggest that impaired OGT/IL-4 regulation occurs within a broader inflammatory shift in cerebellar microglia.

The microglial phenotype identified here should therefore be viewed as one component of a broader developmental effect of *Ogt*^N648Y/y^. OGT controls several pathways directly relevant to cerebellar maturation, including SHH–GLI2 signalling in GCPs^65^, Gα12–RhoA/ROCK signalling in the granule cell lineage^38^ and O-GlcNAc-dependent maintenance of PCs^51^. These observations support a model in which cell-intrinsic neuronal and glial defects are compounded by altered neuroimmune regulation.

In conclusion, our findings establish *Ogt*^N648Y/y^ mice as a clinically relevant model that recapitulates key cerebellar features observed in the affected individual, including impaired postnatal cerebellar growth and associated alterations in cerebellar maturation. Using this model, we identify altered neuroimmune regulation as an important component of the cerebellar pathology associated with the OGT^N648Y^ variant. The developing cerebellum appears particularly dependent on tightly regulated O-GlcNAc signalling, and disruption of this system affects both intrinsic neuronal developmental pathways and IL-4-dependent regulation of microglial state. By linking impaired OGT activity to reduced IL-4 responsiveness, persistent microglial abnormalities and defective postnatal cerebellar maturation, this study broadens the mechanistic framework through which OGT dysfunction may contribute to neurodevelopmental disease and highlights the importance of coordinated neuronal, glial and immune regulation during cerebellar development.

## Data Availability

The bulk RNA-sequencing data generated in this study have been deposited in the NCBI Gene Expression Omnibus (GEO) under accession number GSE347352. Raw clinical neuroimaging data are not publicly available due to patient privacy and ethical restrictions, but relevant derived findings are included in the manuscript. Additional data supporting the findings of this study are available from the corresponding author upon reasonable request.

## Acknowledgments

This work was supported by a Wellcome Trust Investigator Award (110061), Novo Nordisk Foundation Laureate Award (NNF21OC0065969), and Villum Foundation Investigator Award (00054496) to D.M.F.v.A., with additional support from DANDRITE, the Nordic-EMBL Partnership for Molecular Medicine, and the Lundbeck Foundation. Additional funding was provided by the European Regional Development Fund (ERDF) and Fundação para a Ciência e a Tecnologia (FCT; 2022.02604.PTDC and 2023.18329.ICDT). KÕ was supported by Estonian Research grants PRG471 and PRG2040. The authors acknowledge the Microscopy Imaging Center at CNC-UC, the MBG Imaging Facility, and the Bioimaging Core Facility at Aarhus University for access to imaging equipment and technical support. We are grateful to the patient family for their collaboration.

## Author contributions

S.A., S.T.B., J.G., and D.M.F.v.A. conceived the study. S.A., S.T.B., and A.J. performed experiments. L.L. performed bulk RNA sequencing, and L.M. analysed and interpreted the resulting data. G.T. performed immunohistochemistry, and S.T.B. and T.-S.B. performed image analysis. C.L. and J.G. provided training and expertise in imaging and contributed to interpretation of the imaging data. J.P. provided resources and supervision for work performed in his laboratory. M.R.-R. provided resources and supervision for work performed in her laboratory. K.Õ. provided clinical insight and interpreted patient data. P.I. performed MRI analysis and interpreted the imaging findings. S.A., S.T.B., J.G., and D.M.F.v.A. integrated and interpreted the overall study findings and wrote the manuscript with input from all authors.

## Materials and methods

### Reagents and Antibodies

| REAGENT OR RESOURCE | SOURCE | IDENTIFIER |
| --- | --- | --- |
| Recombinant human IL-4 protein | R&D systems | Cat# BT-004 |
| Recombinant mouse IL-4 Protein | R&D systems | Cat# 404-ML |
| OSMI-4 | MedChemExpress | Cat# HY-114361 |
| Thiamet-G | MedChemExpress | Cat# HY-12588 |
| **Antibodies and Stains** |  |  |
| Rabbit anti-MGEA5/OGA (1:1000) | Sigma Aldrich | Cat# HPA036141 |
| Rabbit anti-Arginase-1 (1:1000) | Cell Signaling | Cat# 93668 |
| Mouse anti-OGT (1:1000) | Santa Cruz |  |
| Mouse anti-O-GlcNAc (RL2) (1:500) | Novus Biologicals | Cat# NB300-524 |
| Mouse β-Actin (1:2000) | Merck | Cat# A5441 |
| Mouse IL-4Ra (1:1000) | Santa Cruz | Cat# sc-376253 |
| Rabbit anti-Iba1 (1:800) | Fujifilm Wako | Cat# 019-19741 |
| Goat anti-Iba1 (1:500) | Fujifilm Wako | Cat# 011-27991 |
| Rabbit anti-Calbindin-D-28K (1:1000) | Sigma Aldrich | Cat# C2724 |
| Mouse anti-Calbindin-D-28K (1:1000) | Santa Cruz | Cat #sc-365360 |
| Rabbit anti-Pax6 (1:500) | BioLegend | Cat# 862001 |
| IRDye® 680CW Goat anti-Mouse IgG | LICORbio™ | Cat# 926-32220 |
| IRDye® 680CW Goat anti-Rabbit IgG | LICORbio™ | Cat# 926-32221 |
| IRDye® 800CW Goat anti-Mouse IgG | LICORbio™ | Cat# 926-32210 |
| IRDye® 800CW Goat anti-Rabbit IgG | LICORbio™ | Cat# 925-32211 |
| Alexa Fluor 488 donkey anti-rabbit IgG | Invitrogen | Cat# A32790 |
| Alexa Fluor 488 donkey anti-goat IgG | Invitrogen | Cat# A11055 |
| Alexa Fluor 568 donkey anti-mouse IgG | Invitrogen | Cat# A10037 |
| Alexa Fluor 647 donkey anti-rabbit IgG | Invitrogen | Cat# A31573 |
| 4',6-Diamidine-2'-phenylindole dihydrochloride (DAPI) | Sigma Aldrich | Cat# 28718-90-3 |
| PureBlu™ Hoechst Nuclear Staining Dye | Bio-Rad | Cat# 1351304 |
| **Commercial Kits** |  |  |
| Mouse IL-4 ELISA Kit | Abcam | Cat# AB100710 |
| Pierce™ BCA Protein Assay kit | ThermoFisher Scientific | Cat# 23225 |
| **Experimental Models: Organisms/Strains** |  |  |
| C57BL/6JRj-Ogt<sup>tm1(N648Y)Dva</sup> | Daan van Aalten (AU, Denmark) | N/A |
| HEK-Blue™ IL-4/IL-13 (IL-4) Cells | Invivogen | Cat# hkb-il413-4 |
| **Equipment** |  |  |
| LSM 980 AiryScan 2 Confocal Microscope | Carl Zeiss |  |
| Slide Scanner AxioScan 7 | Carl Zeiss |  |
| Vibratome VT1000S | Leica |  |
| Trans-Blot Turbo Nitrocellulose Packs | Bio-Rad | Cat# 1704159 |
| Precellys 24 Touch Homogenizer | Bertin Technologies | Cat# P002391 |
| **Software and Algorithms** |  |  |
| ZEN Black | Carl Zeiss | RRID:SCR_018163 |
| ZEN Blue | Carl Zeiss | RRID:SCR_013672 |
| Graphpad Prism v10.1.1 | GraphPad Software | https://graphpad.com/ |
| FIJI | Schindelin et al.<sup>66</sup> | https://fiji.sc/ |
| ImageJ | Rueden et al.<sup>67</sup> | https://imagej.net/ |
| QuPath v0.6.0-x64 | Bankhead et al.<sup>68</sup> | https://qupath.github.io/ |

### Animal Husbandry

The *Ogt*^N648Y/y^ line^69^ was maintained on a C57BL/6J background (Janvier, France). Animal cohorts were obtained from crossing of male *Ogt*^+/y^ with female *Ogt*^N648Y/+^ mice. Only male offspring were used for all experimental procedures. The animals were housed in disposable cages with *ad libitum* access to water and food, under 12 h light/dark cycle, at the K40 housing facility of the Department of Molecular Biology and Genetics, Aarhus University. All animal studies and breeding were performed in accordance with the ARRIVE guidelines and the European Communities Council Directive (2010/EU) and were approved by the Danish Animal Experiments Inspectorate, under Breeding license 2022-15-0202-00135 and Project licenses: 2023-15-0201-01426 and 2020-15-0201-00421.

### Microglia isolation and culture

For microglia bulk RNA-seq, P30 male mice from 4 WT and 4 *Ogt*^N648Y/y^ were euthanised by cervical dislocation, brains removed and cerebella dissected. Isolation of microglia was performed by dissociating cerebellar tissue using Adult Brain Dissociation Kit, mouse and rat (# 130-107-677, Miltenyi Biotec) and CD11b (Microglia) MicroBeads, human and mouse (# 130-093-634, Miltenyi Biotec) following the manufacturer’s protocol. For primary microglial cultures, P5 male pups from WT and OGT^N648Y^ were euthanised by decapitation, brains removed and cerebella dissected. Isolation of microglia was performed by dissociating cerebellum tissue using Neural Tissue Dissociation kit (P) (# 130-092-628, Miltenyi Biotec) and CD11b (Microglia) MicroBeads, human and mouse (# 130-093-634, Miltenyi Biotec) following the manufacturer’s protocol. Cells were then seeded onto pre-coated wells with poly-L-lysine (Merck, 15140) in DMEM, 4.5 g/L D-Glucose, L-Glutamine (11965-092, Gibco) supplemented with 10% FBS. Microglia were incubated at 37 °C for 7 days *in vitro* (DIV) prior to assaying with medium change every 2-3 days. Upon DIV7, cultured primary microglia were incubated with either recombinant mouse IL-4 (20 ng/ul) or PBS for 24 h. Post incubation, the cells were harvested for immunoblotting.

### HEK-Blue^TM^ IL-4/IL-13 cells

HEK-Blue IL-4/IL-13 reporter cell lines were purchased from Invivogen (hkb-il413) and cultured in DMEM with 10% Heat Inactivated Fetal Bovine Serum (FBS), 100 U/ml – 100 µg/ml Pen-Strep, 10 µg/ml blasticidin, 100 µg/ml Normocin and 100 µg/ml Zeocin. Cells were seeded into 96-well plates following manufacturer’s recommendations, on the next day cells were incubated with either DMSO, OSMI-4 or Thiamet-G for 2 h prior to incubation with human IL-4 (1 ng/ml) or PBS for 24 h. Medium from the cells was then collected and incubated with QUANTI-Blue™ solution at different timepoints (0.5-3 h) and SEAP activity (absorbance) was measured using a SpectraMax Plus 384 Microplate Reader (Molecular Devices, USA) at 640 nm.

### Immunoblotting

Cells and tissue were harvested in RIPA lysis buffer containing 150 mM NaCl, 1% Nonidet P-40, 0.5% sodium deoxycholate, 0.1% sodium dodecyl sulphate, 25 mM Tris/HCl (pH 7.4), 1 mM sodium orthovanadate, 50 mM sodium fluoride and 5 mM sodium pyrophosphate. Tissue was homogenised in two cycles at 5000 rpm for 30 s with a 10 s break using a Precellys 24 Touch homogeniser (Bertin Technologies). Lysates were centrifuged at 17,000 × g for 10 min at 4 °C, and the protein concentration was determined with Pierce^TM^ BCA Protein Assay kit (ThermoFisher Scientific). Proteins were separated on precast 4-12% NuPAGE Bis–Tris Acrylamide gels (Invitrogen) and transferred to Trans-Blot Turbo Midi 0.2 µm nitrocellulose membranes (Bio-Rad). Membranes were blocked with 5% bovine serum albumin (BSA) in Tris-buffered saline buffer with 0.1% Tween-20 (TBS-T) for 1 h at room temperature (RT) and subsequently incubated with respective primary antibodies in 5% BSA-TBS-T solution overnight at 4 °C. The following day, the membranes were incubated with IRDye^®^ 680/800-labeled secondary IgG antibodies at RT for 1 h. Blots were imaged using a LI-COR Odyssey Infrared Imaging System (LICORbio^TM^) and signals were quantified using Emperia Studio software from the same manufacturer or ImageJ. Antibodies and dilutions are listed under *Reagents and Antibodies*.

### Immunohistochemistry

Mice were transcardially perfused with phosphate-buffered saline (PBS) followed by 4% paraformaldehyde (PFA). Harvested brains were post-fixed overnight at 4 °C in 4% PFA. Brains were then transferred to 15% sucrose in PBS and once submerged, moved to 30% sucrose in PBS containing 0.05% sodium azide for cryoprotection. Samples were kept at 4°C until fully equilibrated, as indicated by loss of buoyancy. Sagittal sections of 50 µm were serially sliced using a vibrating blade microtome (VT1000 S, Leica).

Immunohistochemistry staining was performed on free-floating brain sections^61^. Briefly, the sections were blocked in 5% donkey serum in 0.2% Triton-X-100 in potassium-phosphate buffer (KPBS). The sections were incubated with primary antibodies listed in *Table S1* in the **Supplementary Material**. The following day, sections were washed with KPBS, pre-incubated for 10 min in 1% serum in 0.2% Triton-X-100 in KPBS and incubated for 2 h in the dark with species-specific fluorochrome-conjugated antibodies in 0.2% Triton-X-100 in KPBS with 2.5% serum. Hoechst was added for the last 10 min for nuclear staining. After final washes, sections were mounted on gelatine-coated slides and kept for imaging.

### Fluorescence microscopy imaging and analysis

For histological analysis, stained and mounted sagittal sections were imaged using a Zeiss LSM 980 Confocal Laser Scanning Microscope equipped with an Airyscan 2 detector (Carl Zeiss, Germany, RRID:SCR_018876) and controlled with Zen Black software. A Plan-Apochromat 20×/1.4 NA DIC objective was used for all imaging. For each animal (P10: 5 WT and 8 *Ogt*^N648Y/y^; P30: 6 WT and 5 *Ogt*^N648Y/y^), three cerebellar sagittal sections of sufficient quality were selected. To ensure consistent anatomical coverage across sections, a tiling region was manually delineated around lobule IV/V (vermis) within each section. A 1024 × 1024-pixel Z-stack (20–40 optical planes, 0.6 μm step size) was acquired. Confocal acquisition parameters were kept constant across all slices of the same age and the images were converted to maximum intensity projections and analysed using QuPath open-source software (v.0.6.0). In all microscopy experiments, the experimenter was blind to the genotype, both during acquisition and quantification.

For average layer thickness estimation, three cerebellar lobule IV/V sections were analysed per animal, with three regions of interest (ROIs) delineated in each section. Within each ROI, the cerebellar cortex was subdivided into the molecular layer (ML), Purkinje cell layer (PCL), granular layer (GL), and, for P10 samples, the external granular layer (EGL) was also included. The thickness of each layer was measured in all three ROIs and the measurements were averaged to obtain a mean value for each animal.

For cell density estimation, microglial (IBA1^+^) and Purkinje cell (Calbindin^+^) densities were determined by manually counting marker-positive cell bodies within cerebellar lobules IV/V and dividing the counts by the assessed area (mm²).

Pax6-positive area in lobules IV/V was quantified using ROIs of 9760 µm² for P10 samples and 9563 µm² for P30 samples. Age-specific pixel-based threshold classifiers were generated in QuPath using the AF647-T1 channel at full resolution (0.41 µm/pixel), Gaussian prefiltering, and a smoothing sigma of 1. Intensity thresholds of 41,400 and 22,300 were applied to P10 and P30 sections, respectively. Pixels above the respective threshold were classified as Pax6-positive, whereas pixels below the threshold were classified as negative. Pax6^+^ cell density was expressed as the proportion of Pax6-positive area within each ROI. Measurements from three sections per animal were averaged to obtain a mean value for each animal.

Calbindin expression was obtained using compute intensity features on the ROI with tile size 25 µm. Measurements from three sections per animal were averaged to obtain a mean value for each animal.

For quantification of microglia-mediated phagocytosis, microglia (IBA-1^+^ cells) were probed for the presence of phagocytic cups and counted manually for each image. Measurements from three sections per animal were averaged to obtain a mean value for each animal.

Layer-specific OGT distribution was analysed in P10 sagittal cerebellar sections (n = 3 per genotype). Confocal Z-stacks encompassing cerebellar lobules IV/V were first stitched in ZEN, and the OGT channel was inspected in Fiji to confirm the absence of signal saturation. As the acquired Z-stacks contained different numbers of optical planes, image processing was standardised by selecting a reference plane for each section based on calbindin staining and tissue morphology, independently of the OGT signal. An equal number of Z-planes surrounding the reference plane was then selected for each section, and average-intensity projections were generated for each fluorescence channel.

A fixed-size ROI was applied consistently across all projected images. The ROI was cropped and the individual cerebellar layers were manually delineated using the calbindin channel to avoid bias from the OGT signal. At P10, the EGL, ML, PCL, and IGL were annotated. Mean OGT fluorescence intensity was measured within each layer and corrected by subtraction of the corresponding background fluorescence.

To assess the relative distribution of OGT across the cerebellar cortex, an area-weighted OGT signal was calculated for each layer by multiplying the background-corrected mean OGT fluorescence intensity by the corresponding annotated layer area. The relative contribution of each layer was then calculated as its area-weighted OGT signal divided by the summed area-weighted signal across all layers within the ROI and expressed as a percentage of the total OGT signal. For graphical representation, values were normalised to the mean of the corresponding WT group.

### Enzyme-linked immunosorbent assay (ELISA)

For quantification of IL-4 levels in the parenchyma, ELISA was performed. Anesthetized P10 and P30 mice were perfused with ice-cold PBS and decapitated. The brains were removed from the skulls and dissected into prefrontal cortex, hippocampus and cerebellum. Protein extraction was performed as described for immunoblotting using lysis buffer from the Mouse IL-4 ELISA Kit (Abcam, UK). Subsequently, the Mouse IL-4 ELISA Kit (Abcam, UK) was used according to the manufacturer’s protocol. Briefly, homogenate was diluted 5x with 1× Assay Diluent prior to loading samples into the ELISA microplate, which was incubated overnight at 4 °C in an orbital shaker. The following day, the wells were washed four times with 1× Wash Solution, and 1× Biotinylated Anti-mouse IL-4 Antibody was added and incubated for 1 h at room temperature (RT) with gentle shaking. Unbound antibody was washed away, 1× HRP-Streptavidin Solution was added to bind biotinylated IL-4 antibody and incubated for 45 min at RT with gentle shaking. The wells were washed again, and TMB One-Step Substrate Reagent was added and incubated for 30 min at RT in the dark with gentle shaking to allow colour development (proportional to the amount of bound IL-4). Finally, Stop Solution was added, and the absorbance was measured at 450 nm in a SpectraMax Plus 384 Microplate Reader (Molecular Devices, USA).

### Bulk RNA-sequencing

RNA extraction from isolated microglia was performed using Arcturus^TM^ PicoPure^TM^ RNA isolation kit following the manufacturer protocol. RNA quantity and quality were measured using NanoDrop OneC UV-Vis Spectrophotometer (Thermo Fisher). Polyadenylated mRNAs were reverse transcribed using barcoded primers with UMI and template switching to generate full-length cDNA, which was subsequently amplified. For library construction, amplified cDNA was processed using the Illumina-compatible library preparation workflow with library construction kit (10X Genomics). Final libraries were quality-checked and sequenced on an Illumina NovaSeqX Plus platform. Raw FASTQ files were aligned to the *Mus musculus* reference genome (GRCm39, Ensembl 112) using STAR. Reads were mapped with unique-mapping enabled, unmapped reads retained. Gene-level counts were generated using STAR’s GeneCounts/STARsolo module. UMI sequences were extracted and deduplicated. Alignment statistics and count matrices were produced directly by STAR.

Downstream analyses were performed in R v4.3.3 (Bioconductor 3.18). Genes were restricted to protein-coding genes using the mouse annotation packages org.Mm.eg.db (v3.18.0) and AnnotationDbi (v1.64.1). Low-expression genes were removed by requiring a count of at least [10] in at least [3] samples, leaving ⟨n⟩ genes for analysis. Size-factor-normalised counts were obtained by dividing raw counts by sample-specific size factors estimated with DESeq2::estimateSizeFactorsForMatrix.

Principal component analysis was performed on the filtered counts after size-factor normalisation and log2 transformation (log2[normalised counts + 1]). Per-gene variance was computed across samples and the 2,000 most variable genes retained. Principal components were computed with prcomp on the transposed, scaled expression matrix and visualised with ggplot2 (v4.0.2), with samples coloured by genotype.

Differential expression analysis was performed with DESeq2 (v1.42.1) on the filtered count matrix and matched sample metadata. Counts were rounded to integers, a DESeqDataSet was constructed with DESeqDataSetFromMatrix using the design formula ∼ Genotype, and the model was fitted with DESeq. OgtN648Y/y was compared with wild type with wild type as the reference level. Results were extracted with DESeq2:: results and are reported as base mean expression, log2 fold change, standard error, Wald statistic, raw p value and Benjamini–Hochberg adjusted p value. Differentially expressed genes were defined by an adjusted p value below [0.05]. 189 genes reached nominal p < 0.05 (115 up-regulated and 74 down-regulated in *Ogt*^N648Y/y^ microglia).

For visualisation after model fitting, a variance-stabilising transformation was applied with DESeq2::vst(blind = FALSE). Volcano plots (ggplot2 v4.0.2) display log2 fold change against −log10 nominal p value, with genes classified as up-regulated, down-regulated or not significant using a nominal p cutoff of [0.05] and an absolute log2 fold-change cutoff of [0.5]; the top [20] genes by significance are labelled. Heatmaps (pheatmap v1.0.13) were generated from the variance-stabilised matrix for the top 30 genes ranked by nominal p value, with expression values converted to row-wise Z scores and unsupervised hierarchical clustering of both genes and samples, and a genotype annotation bar.

Gene set enrichment analysis was performed on the full ranked gene list for the OgtN648Y/y versus wild-type contrast. Genes were ranked in decreasing order by the DESeq2 Wald statistic; genes with missing statistics or identifiers were removed and duplicate symbols collapsed, retaining the statistic with the largest absolute value where several symbols mapped to a single Entrez identifier. Gene symbols were mapped to Entrez identifiers with org.Mm.eg.db. Enrichment against Gene Ontology Biological Process terms was computed with clusterProfiler (v4.10.1) using gseGO (organism mmu), with gene-set sizes restricted to between 15 and 500 genes and p values adjusted by the Benjamini–Hochberg method. Terms were ranked by adjusted p value and normalised enrichment score and visualised with enrichplot (v1.22.0) as summary bar plots and running enrichment-score plots. All significantly enriched terms are listed in Supplementary Table 1.

---

**Supplementary Figure 1:**
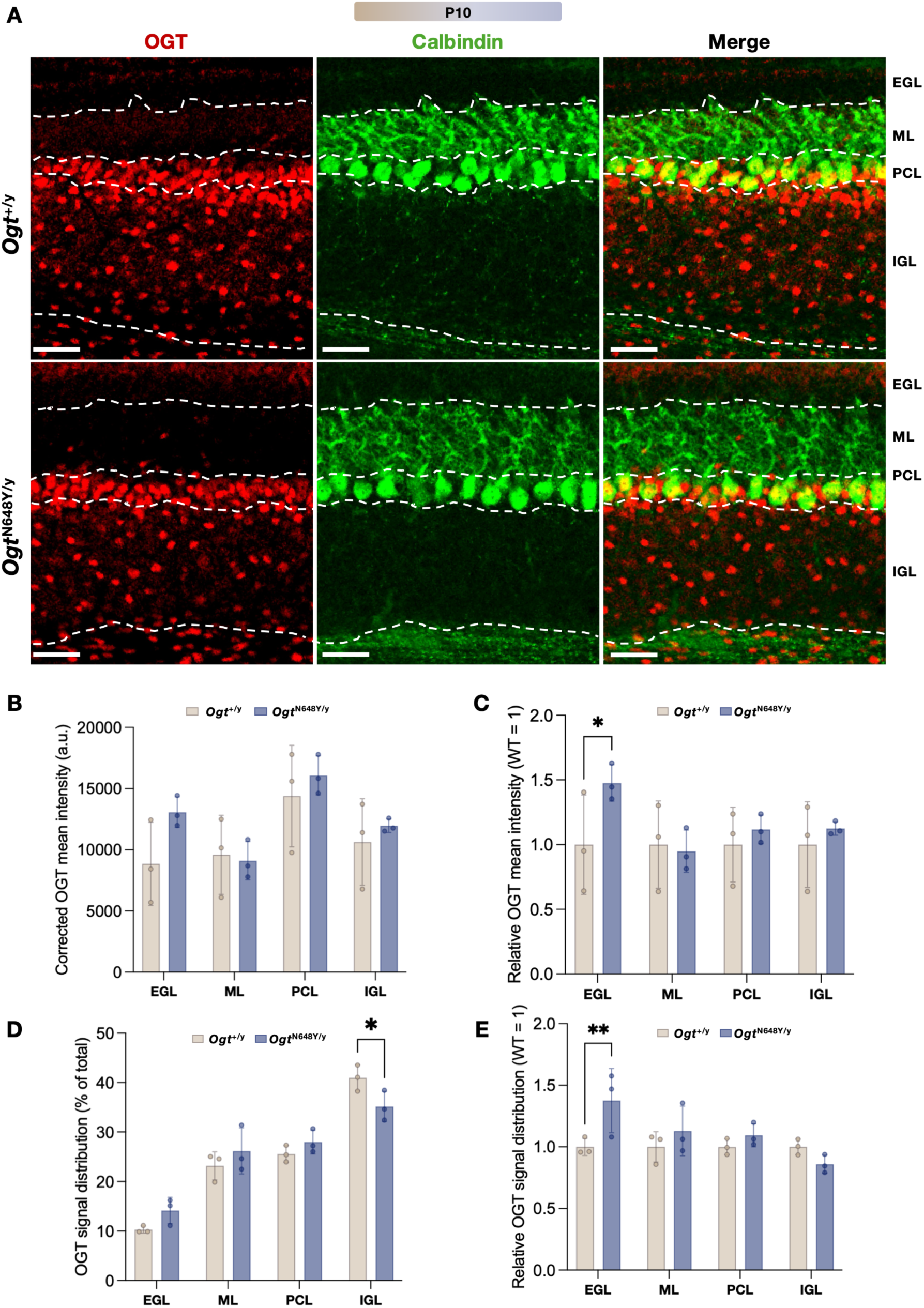
Layer-specific distribution and intensity of OGT signal in the cerebellum. (A) Representative sagittal sections of cerebellar vermis lobules IV/V from WT and *Ogt*^N648Y/y^ mice at P10 immunostained for Calbindin and OGT, illustrating the layer-specific distribution of OGT. Scalebar: 50 µm (B) Area-weighted distribution of OGT signal across the EGL, ML, PCL and IGL, expressed as a percentage of the total OGT signal within the analysed ROI. OGT signal for each layer was calculated as background-corrected mean OGT intensity multiplied by the corresponding layer area. (C) The same area-weighted OGT signal distribution expressed relative to the WT mean for each layer (WT = 1). (D) Background-corrected mean OGT intensity measured within each layer. (E) Background-corrected mean OGT intensity expressed relative to the WT mean (WT = 1). Individual animals are shown as points, with bars representing mean ± SEM. Statistical comparisons are indicated on the graphs. Statistical significance was determined by two-way ANOVA with appropriate multiple-comparison testing; *p < 0.05, **p < 0.01. EGL = external granular layer, ML = molecular layer, PCL = Purkinje cell layer, IGL = internal granule cell layer.

**Supplementary Figure 2:**
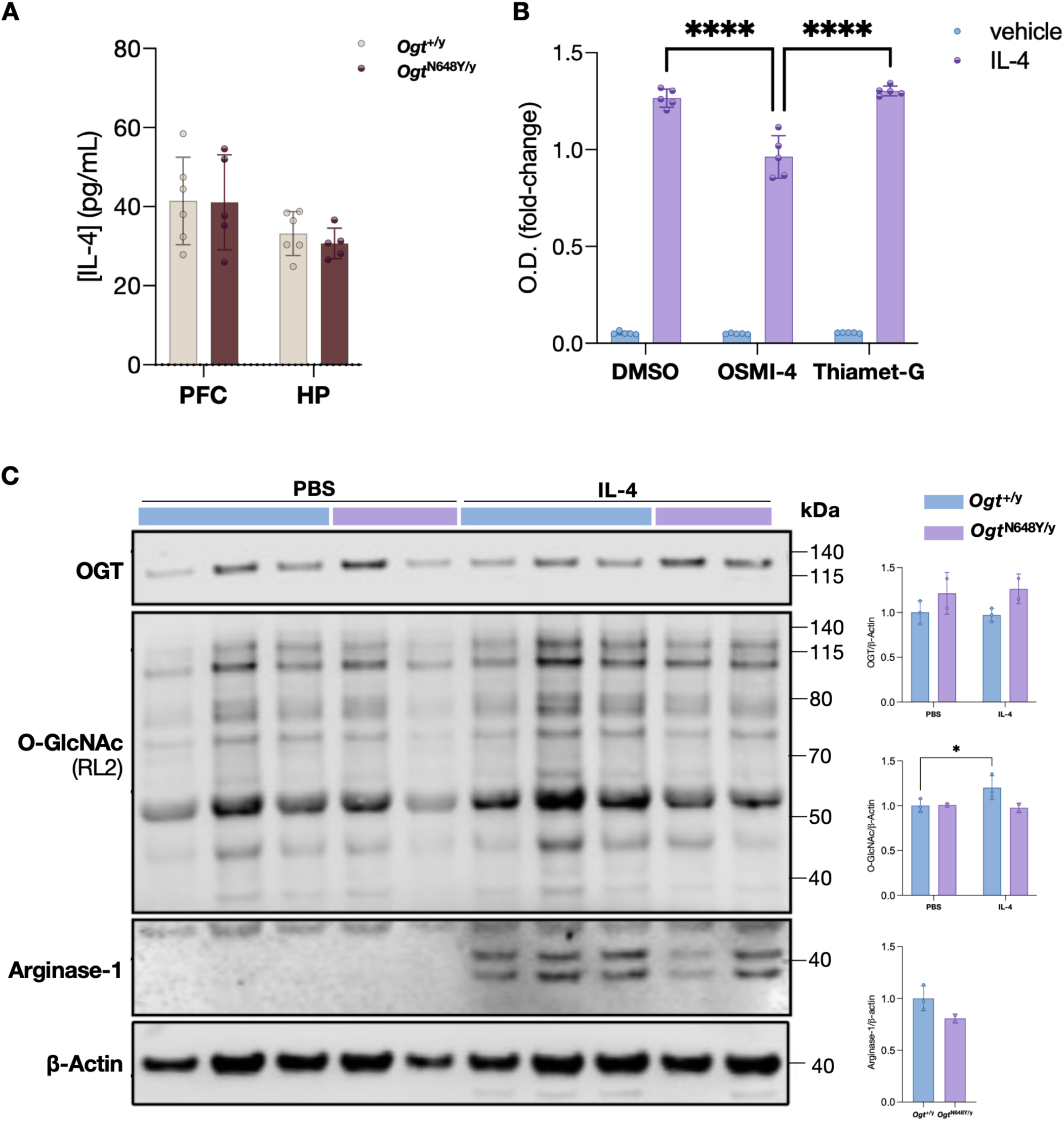
Cerebellum-specific reduction in IL-4 and impaired OGT-dependent IL-4 signalling. (A) IL-4 levels measured by ELISA in of prefrontal cortex and hippocampal lysates at P10 and P30. (B) HEK-Blue IL-4/IL-13 reporter cells were stimulated with IL-4 in the presence of vehicle control (DMSO), OSMI-4 (OGT inhibitor), or Thiamet-G (OGA inhibitor), at 10 µM. SEAP activity was measured as optical density (O.D.) after 30 min as a readout of downstream IL-4/IL-13 signalling output. (C) Western blot analysis and quantification of OGT, global O-GlcNAcylation (RL2) and Arginase-1 in primary microglia isolated from P5 whole brain WT (n=3) and *Ogt*^N648Y/y^ (n=2) mice and stimulated with PBS or IL-4 for 24 h. Relative levels were normalized to β-actin. Data are shown as mean ± SD. Multiple unpaired t-test with Welch’s correction and is used for statistics: *p < 0.05, **p < 0.01, ***p < 0.001.

