## Supplementary figures and images for "Disrupted O-GlcNAc homeostasis links microglial dysfunction to cerebellar hypoplasia"

### Figure 1

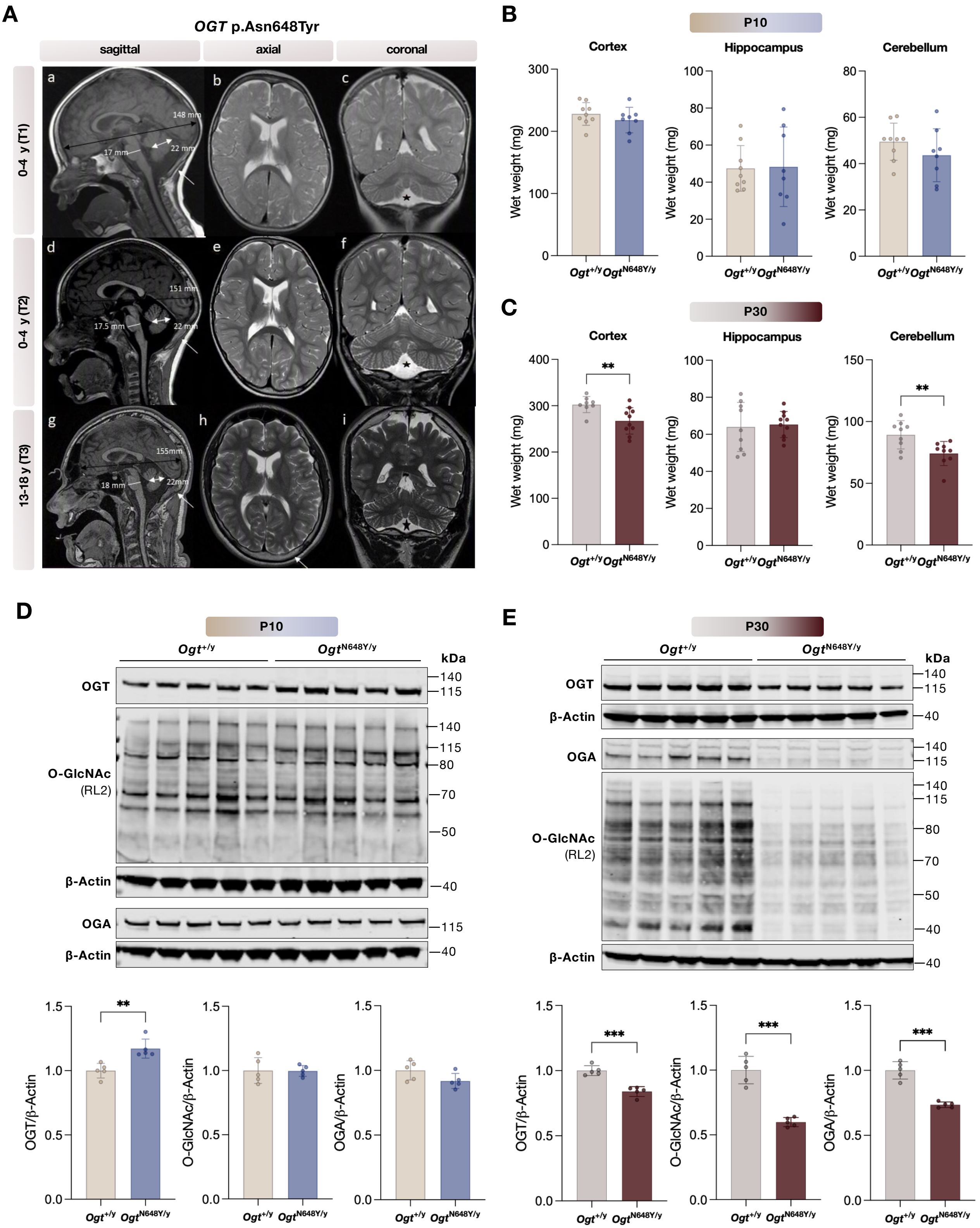

### Figure 2

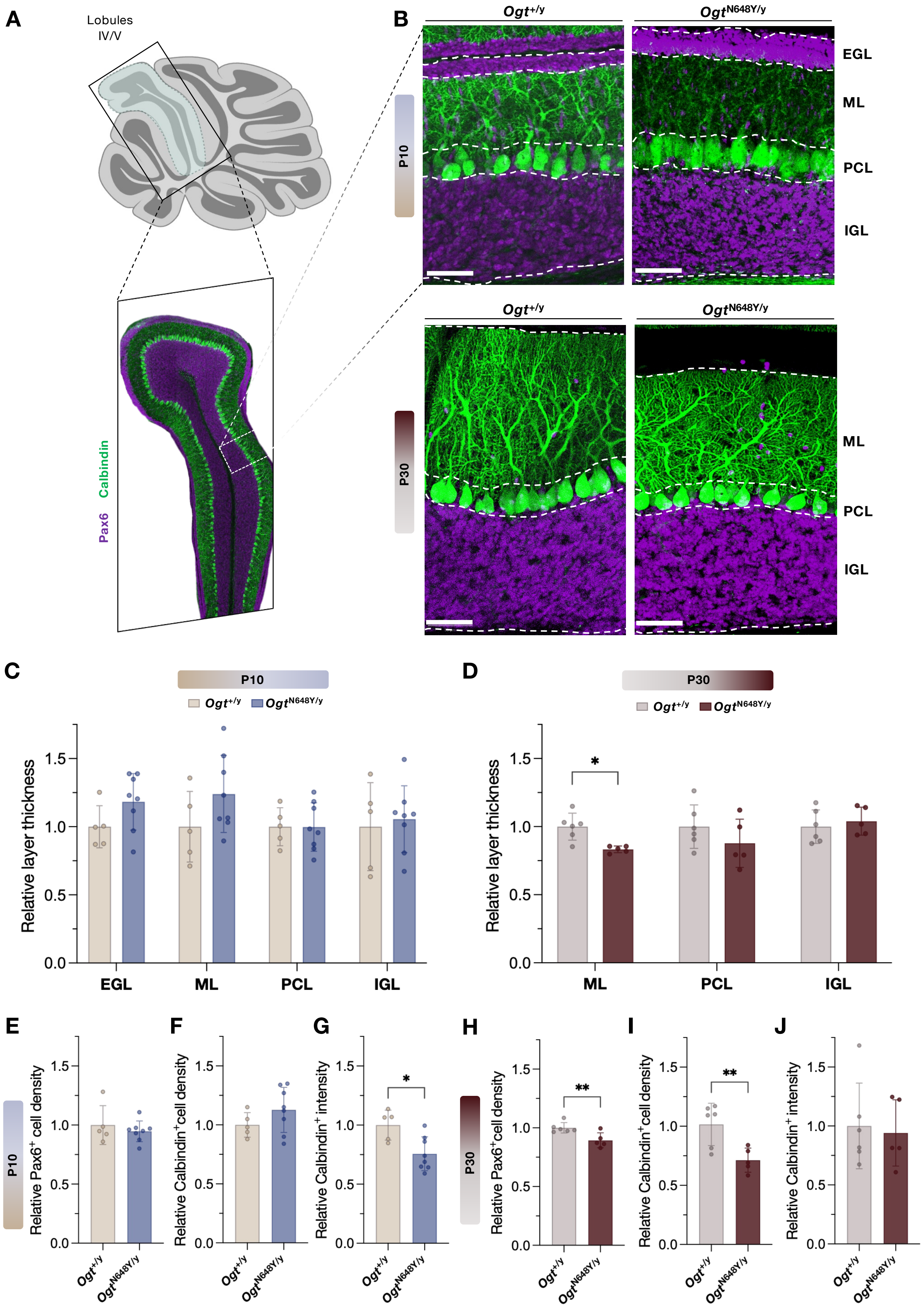

### Figure 3

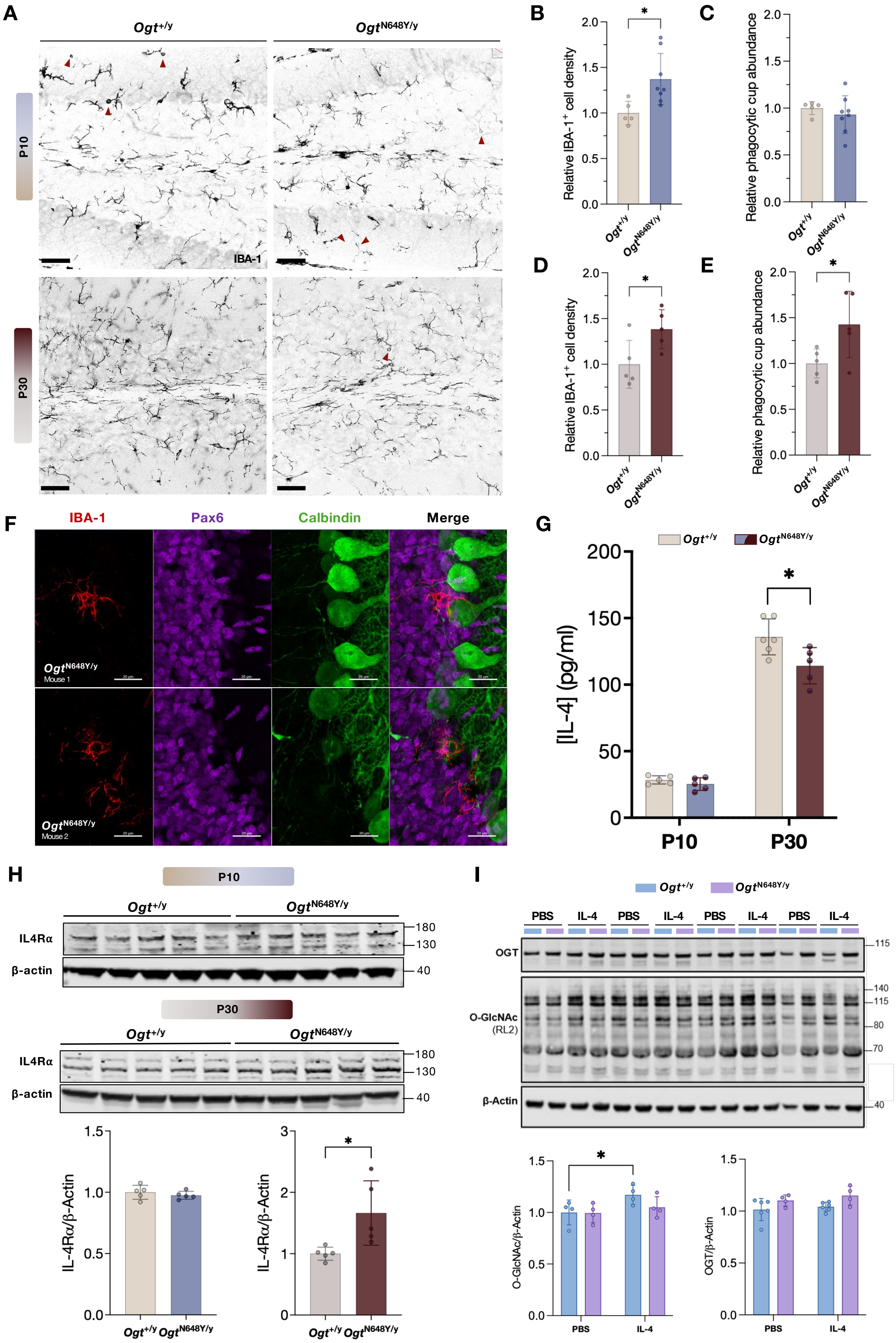

### Figure 4

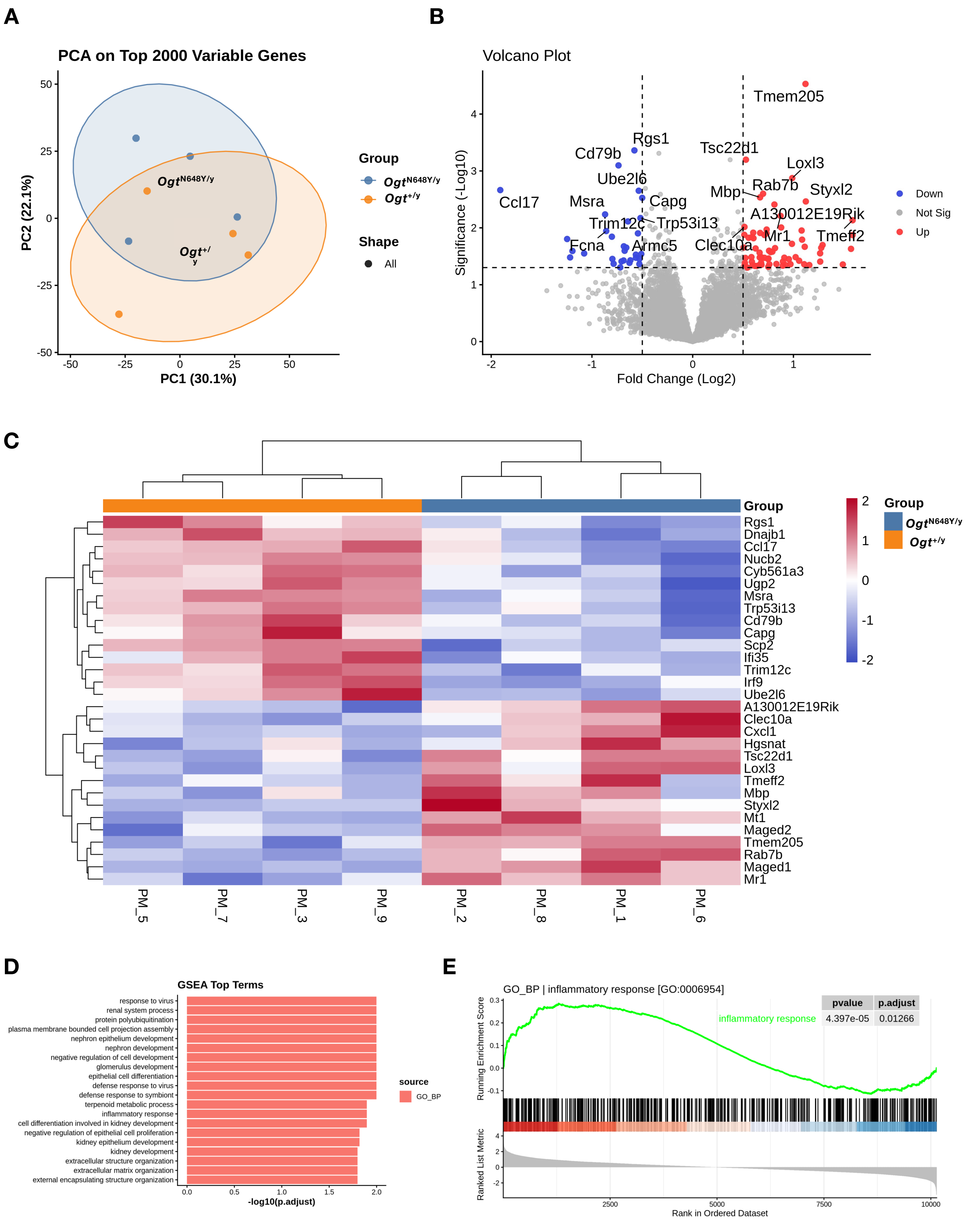

### Sup. Figure 1

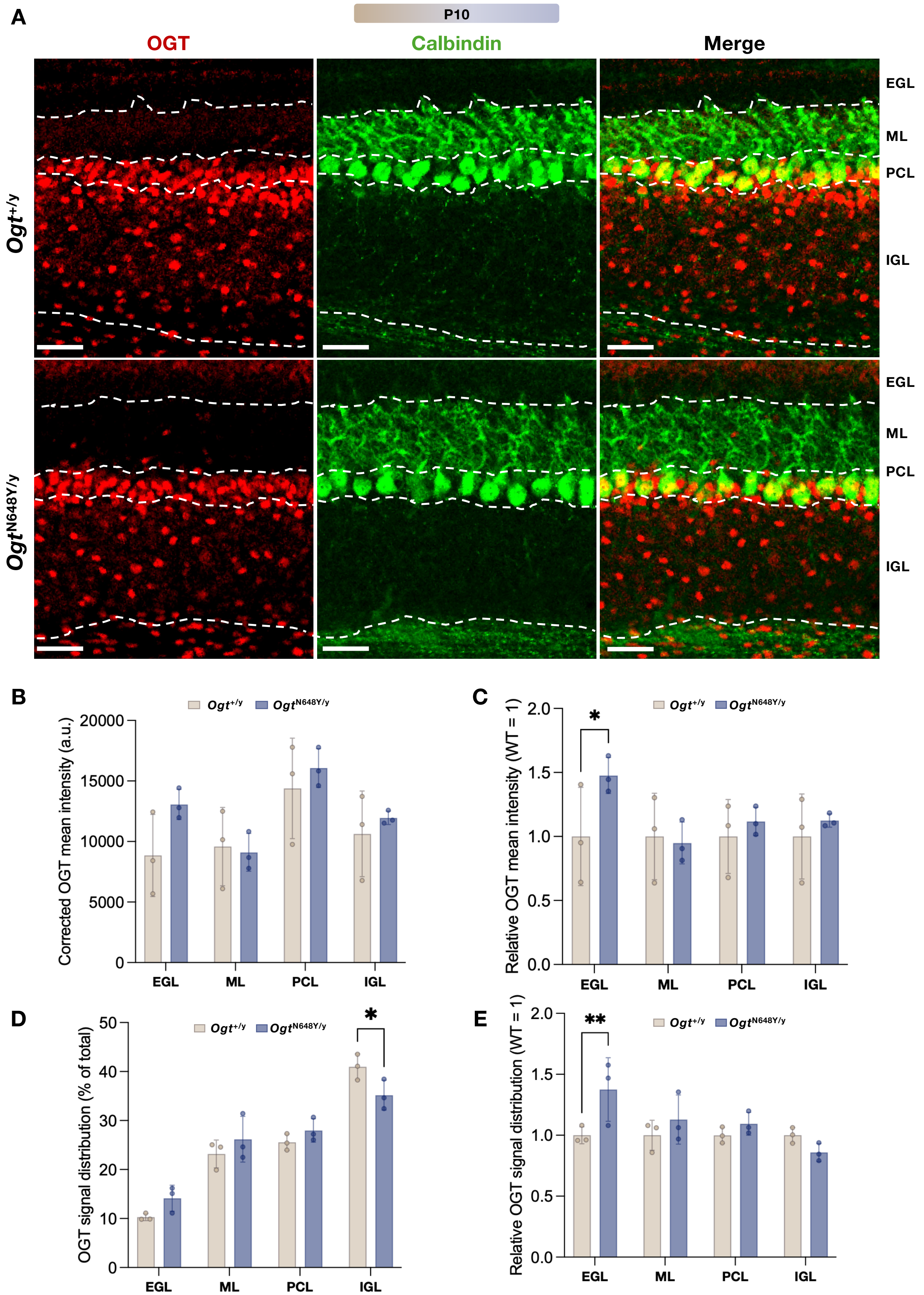

### Sup. Figure 2

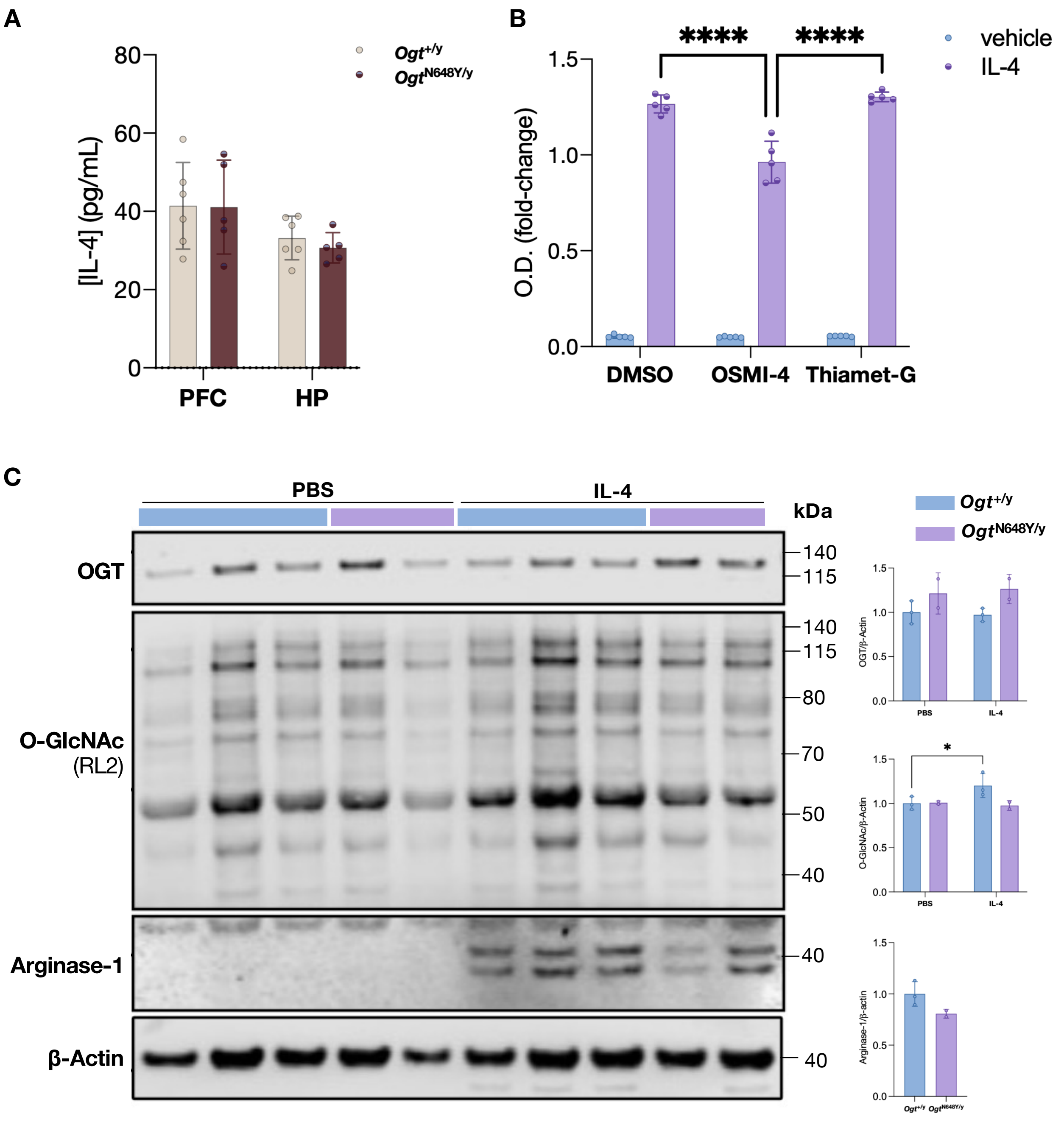
