## Supplementary Table 1 for "Disrupted O-GlcNAc homeostasis links microglial dysfunction to cerebellar hypoplasia"

| Entrez_ID | Gene_symbol | All_symbols | Description |
|---|---|---|---|
| 16950 | Loxl3 | Loxl3, Lor2, Loxl2 | lysyl oxidase homolog 3 precursor |
| 17312 | Clec10a | Clec10a, CD301a, M-ASGP-BP-1, Mgl, Mgl1 | C-type lectin domain family 10 member A isoform 2 |
| 14825 | Cxcl1 | Cxcl1, Fsp, Gro1, KC, Mgsa, N51, Scyb1, gro | growth-regulated alpha protein precursor |
| 12702 | Socs3 | Socs3, Cis3, Cish3, EF-10, Ef10, SSI-3, Ssi3 | suppressor of cytokine signaling 3 |
| 83964 | Jam3 | Jam3, 1110002N23Rik, JAM-3, JAM-C, Jcam3 | junctional adhesion molecule C precursor |
| 20310 | Cxcl2 | Cxcl2, CINC-2a, GROb, Gro2, MIP-2, MIP-2a, Mgsa-b, Mip2, Scyb, Scyb2 | C-X-C motif chemokine 2 precursor |
| 53603 | Tslp | Tslp | thymic stromal lymphopoietin precursor |
| 18858 | Pmp22 | Pmp22, Gas-3, HNPP, PMP-22, TRE002, Tr, trembler | peripheral myelin protein 22 isoform 1 |
| 15894 | Icam1 | Icam1, CD54, Icam-1, Ly-47, MALA-2 | intercellular adhesion molecule 1 precursor |
| 12273 | C5ar1 | C5ar1, C5aR, C5r1, Cd88, D7Msu1 | C5a anaphylatoxin chemotactic receptor 1 |
| 54371 | Chst2 | Chst2, C130041E03Rik, Chts2, GST-2, GlcNAc6ST, Gn6st | carbohydrate sulfotransferase 2 |
| 11816 | Apoe | Apoe, Apo-E | apolipoprotein E precursor |
| 16412 | Itgb1 | Itgb1, 4633401G24Rik, CD29, Fnrb, Gm9863, gpIIa | integrin beta-1 precursor |
| 16176 | Il1b | Il1b, IL-1beta, Il-1b | interleukin-1 beta |
| 19242 | Ptn | Ptn, HARP, HB-GAM, HBBM, HBBN, HBGF-8, HBNF, OSF, Osf-1, Osf1 | pleiotrophin precursor |
| 22695 | Zfp36 | Zfp36, Gos24, Nup475, TIS11D, TISII, Tis11, Ttp, Zfp-36 | mRNA decay activator protein ZFP36 |
| 216799 | Nlrp3 | Nlrp3, AGTAVPRL, AII/AVP, Cias1, FCAS, FCU, MWS, Mmig1, NALP3, Pypaf1 | NACHT, LRR and PYD domains-containing protein 3 |
| 216161 | Sbno2 | Sbno2, F730040C21, Sno, Stno | protein strawberry notch homolog 2 isoform 1 |
| 11990 | Atrn | Atrn, Mgca, mg | attractin preproprotein |
| 384059 | Tlr12 | Tlr12, Gm1365, Tlr11 | toll-like receptor 12 precursor |
| 20303 | Ccl4 | Ccl4, AT744.1, Act-2, MIP-1B, Mip1b, Scya4 | C-C motif chemokine 4 precursor |
| 14815 | Nr3c1 | Nr3c1, GR, Grl-1, Grl1 | glucocorticoid receptor |
| 15945 | Cxcl10 | Cxcl10, C7, CRG-2, INP10, IP-10, IP10, Ifi10, Scyb10, gIP-10, mob-1 | C-X-C motif chemokine 10 precursor |
| 15937 | Ier3 | Ier3, IEX-1, cI-3, gly96 | radiation-inducible immediate-early gene IEX-1 |
| 16852 | Lgals1 | Lgals1, Gal-1, Galbp, L-14.5, L14, Lect14 | galectin-1 |
| 56744 | Pf4 | Pf4, Cxcl4, Scyb4 | platelet factor 4 precursor |
| 15289 | Hmgb1 | Hmgb1, HMG-1, Hmg1, SBP-1, p30 | high mobility group protein B1 |
| 24136 | Zeb2 | Zeb2, 9130203F04Rik, D130016B08Rik, SIP1, Zfhx1b, Zfx1b, Zfxh1b | zinc finger E-box-binding homeobox 2 |
| 244329 | Mcph1 | Mcph1, 5430437K10Rik, BRIT1, D030046N04Rik, MCT, Tg(HLA-A2.1)1Enge | microcephalin isoform 3 |
| 53416 | Stk39 | Stk39, DCHT, Gm50618, RF005, Rnl5, SPAK | STE20/SPS1-related proline-alanine-rich protein kinase isoform 1 |
| 73242 | Atat1 | Atat1, 0610011P08Rik, 2610008K08Rik, 2610110G12Rik, 3110080J08Rik, MEC-17, Mec17, TAT | alpha-tubulin N-acetyltransferase 1 isoform 1 |
| 14609 | Gja1 | Gja1, Cnx43, Cx43, Cx43alpha1, Cxnk1, Gja-1, Npm1, connexin43 | gap junction alpha-1 protein |
| 18787 | Serpine1 | Serpine1, PAI-1, PAI1, Planh1 | plasminogen activator inhibitor 1 precursor |
| 19219 | Ptger4 | Ptger4, EP4, Ptgerep4 | prostaglandin E2 receptor EP4 subtype isoform 2 |
| 13032 | Ctsc | Ctsc, CatC, DPP1, DPPI | dipeptidyl peptidase 1 isoform 1 preproprotein |
| 56312 | Nupr1 | Nupr1, 2310032H04Rik, Com1, p8 | nuclear protein 1 |
| 16175 | Il1a | Il1a, Il-1a | interleukin-1 alpha |
| 26362 | Axl | Axl, Ark, Tyro7, Ufo | tyrosine-protein kinase receptor UFO isoform 1 precursor |
| 22177 | Tyrobp | Tyrobp, DAP12, KARAP, Ly83 | TYRO protein tyrosine kinase-binding protein precursor |
| 16784 | Lamp2 | Lamp2, CD107b, LGP-B, Lamp II, Lamp-2, Lamp-2a, Lamp-2b, Lamp-2c, Mac3 | lysosome-associated membrane glycoprotein 2 isoform 1 precursor |
| 18035 | Nfkbia | Nfkbia, Nfkbi | NF-kappa-B inhibitor alpha |
| 16153 | Il10 | Il10, CSIF, If2a, Il-10 | interleukin-10 precursor |
| 14127 | Fcer1g | Fcer1g, CD23, FcR-gamma, FcR[g], FcRgamma, Fce1g, FcepsilonRI, Ly-50 | high affinity immunoglobulin epsilon receptor subunit gamma precursor |
| 16000 | Igf1 | Igf1, C730016P09Rik, Igf-1, Igf-I | insulin-like growth factor I isoform 4 preproprotein |
| 18128 | Notch1 | Notch1, 9930111A19Rik, Mis6, N1, Tan1, lin-12 | neurogenic locus notch homolog protein 1 precursor |
| 57261 | Brd4 | Brd4, Brd5, HUNK1, MCAP, WI-11513 | bromodomain-containing protein 4 isoform 1 |
| 20296 | Ccl2 | Ccl2, HC11, JE, MCAF, MCP-1, MCP1, SMC-CF, Scya2, Sigje | C-C motif chemokine 2 precursor |
| 18823 | Plp1 | Plp1, DM20, Plp, jimpy, jp, msd, rsh | myelin proteolipid protein isoform 1 |
| 14066 | F3 | F3, CD142, Cf-3, Cf3, TF | tissue factor precursor |
| 56644 | Clec7a | Clec7a, BGR, Clecsf12, beta-GR | C-type lectin domain family 7 member A isoform 2 |
| 19204 | Ptafr | Ptafr, PAFR | platelet-activating factor receptor |
| 13618 | Ednrb | Ednrb, ET-B, ET-BR, ETR-b, ETb, Sox10m1 | endothelin receptor type B isoform 1 precursor |
| 12759 | Clu | Clu, ApoJ, Cli, D14Ucla3, SP-40, Sgp-2, Sgp2, Sugp-2 | clusterin preproprotein |
| 67389 | C1qtnf12 | C1qtnf12, 1110035L05Rik, C1qdc2, CTRP12, Fam132a, alipolin | adipolin precursor |
| 14775 | Gpx1 | Gpx1, CGPx, GPx-1, GSHPx-1, Gpx | glutathione peroxidase 1 isoform 1 |
| 338364 | Trim65 | Trim65, 4732463G12Rik | E3 ubiquitin-protein ligase TRIM65 |
| 14131 | Fcgr3 | Fcgr3, CD16 | low affinity immunoglobulin gamma Fc region receptor III isoform 2 precursor |
| 14154 | Fem1a | Fem1a, Eprap, Fem1aa | protein fem-1 homolog A-A |
| 72565 | Uaca | Uaca, 2700059D02Rik, nucling | uveal autoantigen with coiled-coil domains and ankyrin repeats isoform 2 |
| 19261 | Sirpa | Sirpa, Bit, CD172a, Idd13.2, P84, Ptpns1, SHP-1, SHPS-1, SIRP | tyrosine-protein phosphatase non-receptor type substrate 1 isoform 3 precursor |
| 24088 | Tlr2 | Tlr2, Ly105 | toll-like receptor 2 precursor |
| 18750 | Prkca | Prkca, Pkca | protein kinase C alpha type |
| 20306 | Ccl7 | Ccl7, MCP-3, Scya7, fic, marc, mcp3 | C-C motif chemokine 7 precursor |
| 21926 | Tnf | Tnf, DIF, TNF-a, TNF-alpha, TNFSF2, TNFalpha, Tnfa, Tnfsf1a, Tnlg1f | tumor necrosis factor isoform 1 |
| 12267 | C3ar1 | C3ar1, AZ3B, C3AR, HNFAG09 | C3a anaphylatoxin chemotactic receptor |
| 319430 | C5ar2 | C5ar2, C5L2, E030029A11Rik, Gpr77 | C5a anaphylatoxin chemotactic receptor 2 isoform 2 |
| 19696 | Rel | Rel, c-Rel | proto-oncogene c-Rel |
| 12774 | Ccr5 | Ccr5, AM4-7, CD195, Cmkbr5 | C-C chemokine receptor type 5 |
| 20300 | Ccl25 | Ccl25, A130072A22Rik, CKb15, Scya25, TECK | C-C motif chemokine 25 precursor |
| 11491 | Adam17 | Adam17, CD156b, Tace | disintegrin and metalloproteinase domain-containing protein 17 isoform 1 preproprotein |
| 13011 | Cst7 | Cst7, Cmap | cystatin-F precursor |
| 230738 | Zc3h12a | Zc3h12a, MCPIP, MCPIP-1, Mcpip1, Reg1 | endoribonuclease ZC3H12A |
| 63873 | Trpv4 | Trpv4, 0610033B08Rik, OTRPC4, Trp12, VR-OAC, VRL-2, VROAC | transient receptor potential cation channel subfamily V member 4 |
| 17096 | Lyn | Lyn, Hck-2, p53Lyn, p56Lyn | tyrosine-protein kinase Lyn isoform A |
| 17079 | Cd180 | Cd180, F630107B15, Ly-78, Ly78, RP105 | CD180 antigen isoform 1 precursor |
| 21803 | Tgfb1 | Tgfb1, TGF-beta1, TGFbeta1, Tgfb, Tgfb-1 | transforming growth factor beta-1 proprotein preproprotein |
| 97165 | Hmgb2 | Hmgb2, HMG-2, Hmg2 | high mobility group protein B2 |
| 216850 | Kdm6b | Kdm6b, 1700064E03Rik, Jmjd3 | lysine-specific demethylase 6B |
| 18807 | Pld3 | Pld3, Sam-9 | 5'-3' exonuclease PLD3 isoform a |
| 18129 | Notch2 | Notch2, N2 | neurogenic locus notch homolog protein 2 precursor |
| 80859 | Nfkbiz | Nfkbiz, INAP, Mail | NF-kappa-B inhibitor zeta isoform a |
| 18034 | Nfkb2 | Nfkb2, NF-kappaB2, lyt, p49, p49/p100, p50B, p52 | nuclear factor NF-kappa-B p100 subunit isoform a |
| 16803 | Lbp | Lbp, Bpifd2, Ly88 | lipopolysaccharide-binding protein precursor |
| 74748 | Slamf8 | Slamf8, 5830408F06Rik, Blame, SBBI42 | SLAM family member 8 precursor |
| 20655 | Sod1 | Sod1, B430204E11Rik, Cu/Zn-SOD, CuZnSOD, Ipo-1, Ipo1, SODC, Sod-1 | superoxide dismutase [Cu-Zn] |
| 54131 | Irf3 | Irf3, C920001K05Rik, IRF-3 | interferon regulatory factor 3 |
| 15467 | Eif2ak1 | Eif2ak1, HCR, Hri | eukaryotic translation initiation factor 2-alpha kinase 1 isoform 1 |
| 12266 | C3 | C3, ASP, HSE-MSF, Plp | complement C3 preproprotein |
| 69769 | Tnfaip8l2 | Tnfaip8l2, 1810019A08Rik, TIPE2 | tumor necrosis factor alpha-induced protein 8-like protein 2 |
| 64297 | Gprc5b | Gprc5b, Raig2 | G-protein coupled receptor family C group 5 member B isoform 2 precursor |
| 12475 | Cd14 | Cd14 | monocyte differentiation antigen CD14 precursor |
| 26874 | Abcd2 | Abcd2, ABC39, ALDL1, ALDR, ALDRP | ATP-binding cassette sub-family D member 2 isoform 1 |
| 279572 | Tlr13 | Tlr13, Gm713 | toll-like receptor 13 precursor |
| 56221 | Ccl24 | Ccl24, CKb-6, MPIF-2, Scya24 | C-C motif chemokine 24 precursor |
| 14000 | Drosha | Drosha, 1110013A17Rik, Etohi2, Rn3, Rnasen | ribonuclease 3 |
